# CovidSIMVL --Transmission Trees, Superspreaders and Contact Tracing in Agent Based Models of Covid-19

**DOI:** 10.1101/2020.12.21.20248673

**Authors:** Ernie Chang, Kenneth A. Moselle, Ashlin Richardson

## Abstract

The agent-based model CovidSIMVL (github.com/ecsendmail/MultiverseContagion) is employed in this paper to delineate different network structures of transmission chains in simulated COVID-19 epidemics, where initial parameters are set to approximate spread from a single transmission source, and R_0_ranges between 1.5 and 2.5.

The resulting Transmission Trees are characterized by breadth, depth and generations needed to reach a target of 50% infected from a starting population of 100, or self-extinction prior to reaching that target. Metrics reflecting efficiency of an epidemic relate closely to topology of the trees.

It can be shown that the notion of superspreading individuals may be a statistical artefact of Transmission Tree growth, while superspreader events can be readily simulated with appropriate parameter settings. The potential use of contact tracing data to identify chain length and shared paths is explored as a measure of epidemic progression. This characterization of epidemics in terms of topological characteristics of Transmission Trees may complement equation-based models that work from rates of infection. By constructing measures of efficiency of spread based on Transmission Tree topology and distribution, rather than rates of infection over time, the agent-based approach may provide a method to characterize and project risks associated with collections of transmission events, most notably at relatively early epidemic stages, when rates are low and equation-based approaches are challenged in their capacity to describe or predict.

**MOTIVATION – MODELS KEYED TO CONTEMPLATED DECISIONS:** Outcomes are altered by changing the processes that determine them. If we wish to alter contagion-based spread of infection as reflected in curves that characterize changes in transmission rates over time, we must intervene at the level of the processes that are directly involved in preventing viral spread. If we are going to employ *models* to evaluate different candidate arrays of localized preventive policies, those models must be posed at the same level of *granularity* as the entities (people enacting processes) to which preventive measures will be applied. As well, the models must be able to represent the transmission-relevant *dynamics* of the systems to which policies could be applied. Further, the *parameters* that govern dynamics within the models must embody the actions that are prescribed/proscribed by the preventive measures that are contemplated. If all of those conditions are met, then at a formal or structural level, the models are conformant with the provisions of the Law of Requisite Variety^1^ or the restated version of that law – the good regulator theorem.^2^

On a more logistical or practical level, the models must yield *summary measures* that are responsive to changes in key parameters, highlight the dynamics, quantify outcomes associated with the dynamics, and communicate that information in a form that can be understood correctly by parties who are adjudicating on policy options.

If the models meet formal/structural requirements regarding requisite variety, and the parameters have a plausible interpretation in relationship to real-world situations, and the metrics do not overly-distort the data contents that they summarize, then the models provide information that is directly relevant to decision-making processes. Models that meet these requirements will minimize the gap that separates models from decisions, a gap that will otherwise be filled by considerations other than the data used to create the models (for equation-based models) or the data generated by the simulations.

In this work, we present an agent-based model that targets information requirements of decision-makers who are setting policy at a local level, or translate population level directives to local entities and operations. We employ an agent-based modeling approach, which enables us to generate simulations that respond directly to the requirements of the good regulator theorem. Transmission events take place within a spatio-temporal frame of reference in this model, and rates are not conditioned by a reproduction rate (R0) that is specified *a priori*. Events are a function of movement and proximity. To summarize dynamics and associated outcomes of simulated epidemics, we employ metrics reflecting topological structure of transmission chains, and distributions of those structures. These measures point directly to dynamic features of simulated outbreaks, they operationalize the “efficiency” construct, and they are responsive to changes in parameters that govern dynamics of the simulations.

## BACKGROUND^3^

### What is CovidSIMVL, and in what are contexts may it be a useful tool?

CovidSIMVL is short-hand for “Covid-19 Simulation, Viral Load version”. It is an agent-based infectious disease modeling tool used to simulate viral transmission within a given local spatial context-- or within a functionally interconnected array of local spatial contexts.

Agents are deemed susceptible, incubating, infective or inert at each simulation trial iteration and are located in finite spatial contexts or interconnected arrays thereof. Upon initialization, parameters are set to capture proximity and movement, to reflect spatial and temporal infection spread features observed in real world contexts. Parameters can be modified to embody various protections e.g., masking.

Over the course of iterations within a CovidSIMVL trial, agents move stochastically within and between interconnected spaces, giving rise to Transmission Trees of varying structure for a given set of initialization parameters. At each iteration in a simulation trial, transmission events ensue, reflecting pathophysiology and viral transmission mechanisms, density of agents within local contexts, and movement within and between contexts.

Topology of Transmission Trees and associated distribution of branches of different lengths can be interpreted directly in terms of the construct “dispersion” (i.e., dispersion parameter *k*) ^4^ which is typically invoked to describe features of outbreaks where variation in infectiousness among individuals is materially consequential with regard to dynamic features of outbreaks. These features include the likely extent of spread, the speed at which a an infection is likely to propagate out through a population, and the risk for explosive outbreaks that represent sharp departures from prevailing population growth rates and are not reflected in the typically smooth curves that are fit to population-level data in typical equation-based models.^5,6^ Modeling of non-linear or localized features of outbreak *via* analysis of the topology of Transmission Trees is relevant to decision-making around individual-specific or context-specific control measures.

### Compartmental Equation-based models – fitting mathematical models to events (e.g., new infections)

Equation-based compartmental models of epidemic spread fit a system of equations to a set of historical data e.g., to new infections, proportion of test positives, or deaths. The data points reflect the outcomes of processes from which new infections arise. Future states are predicted by extrapolating from equations derived from historical data. Validity of those predictions rests on assuming that there is a pattern of transmission dynamics underlying the historical observations which will persist into the future, or which will be reproduced in some new setting to which the equations-based model is applied.

Equation-based compartmental models typically operate on the “mass action incidence” principle^5^:

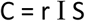

where C is the rate of new infections, r is a constant, I is the number of infectious individuals, S is the number of susceptible individuals. This assumes individuals within a population are evenly mixed i.e. all contacts are equally probable per unit time, thus probabilities of transmission from infected to susceptible are identical per unit time.^7^

Transmission dynamics heterogeneity (reflected, for example, in different reproduction rates) or context-bound population vulnerabilities can be accommodated by homogeneously partitioning suitably large populations into local but still large sub-population contexts – “patches”^8^ which can be used to represent homes and home-based transmission, schools, long-term care facilities, or other local contexts. These are assembled into “meta-population” variants of the equation-based models.^9^

Computational and sample size challenges may be associated with meta-population partitioning of real-world datasets to represent relevant local transmission contexts.^10^ Also, when degree of infectiousness arises from complex interactions of host behaviour, pathogen and environmental factors, it may be impossible to define risk groups on *a priori* grounds,^11^ or datasets may not supply sufficient information required to partition data to reflect real-world assembly of contexts into a network conformant to intra-contextual movement of persons.^12^

### Agent-based models – generating events

Compartmental models of viral transmission fit systems of differential equations to health events that occur over time. Agent-based models iteratively generate discrete events, the properties of which may not always be well-represented or readily reproduced by systems of ordinary differential equations.^13^ These events *are* intended to embody realistic transmissions in arrays of spatio-temporal contexts where distinct transmission dynamics are supported in each context, such as a long-term care facility consisting of a mixture of staff and residents *vs* a very different maximum 23 hour staffed shelter for substance-using individuals, who spend the remainder of their 24 hour days in locations where they could become infected or transmit infection, such as the streets, other sheltered environments, or various other services for persons experiencing homelessness. CovidSIMVL’s flexibility to represent realistic possibilities is achieved *via* parameters governing agent behaviour that varies as a function of location and agent movement and is subject to rules reflecting transmission pathophysiology. Simulation events or averaged quantities reflecting them may indicate significant transmission variation associated with distinguishable localized subgroups contained in larger susceptible populations.

### Agent-based models – characterizing events

Like other agent-based or individual-based infectious disease transmission models, ^14,15,16^ CovidSIMVL generates aggregate results by counting events over the course of multiple steps within a given simulation trial. These aggregate results include regularly reported metrics such at R_0_ at a particular simulation step. Similar rates of infectiousness across contexts and iterations within a trial may suggest invariance with respect to model parameter value assignments e.g., duration of infectivity directly related to properties of the infectious organism. In setting the parameters to produce results of this nature, such results may sometimes be meaningfully summarized by parametric curves associated with systems of ordinary differential equations – that is, the simulation may generate results consistent with models governed by the “mass action incidence” principle.

However, significant transmission variation (e.g., “superspreading events”) is usual for infectious disease spread.^17,18,19^ This variability is provided with every “opportunity” to manifest itself in an agent-based model. Specifically, by tracking direct linkages between events and assembling them into Transmission Trees, we create measures of transmission dynamics reflecting topological transmission attributes which may represent infection vitality in a way that is not necessarily captured in smooth curves generated by equation-based models.

Because CovidSIMVL agent-based simulations evolve from properties of individual agents who are not *a priori* inherently mutually differentiated, heterogeneity for agent-based models can be reflected directly by initial agent positions. Parameter settings capture transmission-relevant properties. Consequently, in modeling scenarios where heterogeneity is required based on “prior” knowledge of distinctive characteristics of local settings, and stochastic and spatial effects contribute to system dynamics,^20^ agent-based models are suited to the task.

### Transmission Chains, Transmission Trees and Transmission Forests

A ***Transmission Chain*** C is a finite sequence C_n_ of individuals where the first element of the sequence C_0_ an infectious agent, and for all applicable n we have viral transmission events between consecutive agents i.e. C_n_ infects C_n+1_. So C traces transmission events along a path. For convenience we state C to be rooted at C_0_. A ***Transmission Tree T*** *(rooted at T*_*0*_ where T_0_ is an initially infectious individual) is a directed graph (V,E) where the node set V is comprised of T_0_ plus any individuals belonging to Transmission Chains such that C_0_ = T_0_. Then the directed edge set E consists of all ordered pairs (C_n_, C_n+1_) comprised of consecutive agents belonging to any Transmission Chain corresponding to Transmission Chains C such that C_0_ = T_0_. Then the degree of a node is the number of individuals subsequently infected by that node. Finally, a ***Transmission Forest*** is a set of Transmission Trees with distinct roots.

Since CovidSIMVL tracks transmission events C_n_ infects C_n+1_ for all agents at each step of a given simulation trial, the tool characterizes outbreaks as sets of Transmission Chains, relating the progression within a single array or a heterogeneous array of local transmission contexts, to the emergence of topologically distinct networked chains of varying lengths. These chains determine a rooted Transmission Tree for a simulation trial seeded with a single infective agent, or determine a Transmission Forest should the simulation be primed with multiple infective agents.

This process of constructing Transmission Trees from CovidSIMVL simulations differs from the inferred tree structures derived from epidemiological data consisting of new case dates and location within geographical division that may or may not map directly or cleanly onto the spaces within which transmission events occurs. These trees are statistical reconstructions whose validity reflects level of detail in contact information.^21^ In CovidSIMVL Transmission Trees are encoded directly from an event log generated over the course of a simulation trial. No inference is involved in constructing these trees.

### Fit-for-purpose modeling

Models should be fit for the purpose of adapting to intended scope and scale of phenomena, including scope and scale of decisions to be supported or rationalized by the models. Equation-based models describing populations holistically and treating population members as functionally identical may be most transparently relevant and interpretable when contemplating policies to be applied at population level, where stochasticity and heterogeneity at the level of local transmission may not be relevant. Use of equation-based models at this scale of policy setting may conform well to the Law of Requisite Variety, and the models and metrics may provide best possible evidence to support decision making.

Agent-based models may more flexibly illuminate early stages of infection transmission, when rates are low and infection has not dispersed throughout communities, where stochasticity is an important factor in determine breadth and level of spread, or when local context-specific decision supports are needed e.g., in considering targeted selective restart of ambulatory services to manage risk for healthcare associated transmission, or in considering operational continuity for short-term emergency shelters while possibly incurring cross-over transmission risk for populations experiencing homelessness.

Regarding compartmental equation-based vs agent-based models, we need not consider either exclusively. A “most-transparently useful and demonstrably valid” issue may warrant different approaches given different envisioned informational uses moreover agent-based models and equation-based models are treated as complementary, as reflected in the two modeling streams promoted by organizations such as the Public Health Agency of Canada.^22^ Further, hybrid models fully integrating both approaches are developed, and are demonstrably useful approaches to overcoming limitations inherent in reliance on one or the other approach.^23^ The work presented in this paper may be considered to be preparatory to such an approach.

### Single universe, multiverse implementation of CovdSIMVL

CovidSIMVL can be configured to simulate spread among a prescribed number of susceptible persons occupying a single physically delimited spatial context (“Universe”). In a previous paper, simulations within these single Universes calibrate key CovidSIMVL parameters against growth curves associated with different R_0_ values^24^. The same single-Universe version of CovidSIMVL generates the Transmission Chains examined in this manuscript.

The Multiverse version of CovidSIMVL simulates epidemics in arrays of local spatial contexts that may be heterogeneous with respect to parameters determining rate and extent of spread. This Multiverse version consists of up to nine CovidSIMVL Universes (e.g., school, homes, restaurants, long-term care facilities) traversed by agents according to schedules reflecting patterned movements over the course of weekly schedules. As individual agents can move between Universes, this CovidSIMVL version accommodates arrays of agents of differing infectiousness, where the degree of infectiousness for an agent is assumed to modulate with agent location within a given Universe. This version supplies the transmission-event-based Transmission Chains and other metrics discussed in Chang, Moselle & Richardson, 2020 (in preparation).^25^

### Configuration

CovidSIMVL enables model calibration by employing rules organized in a three-fold hierarchy to support the injection of biological, behavioural and local spatially-contextual heterogeneity that we deem to be logically necessary to support a diversity of realistic model scenarios, as per conditions set by the “Law of Requisite Variety” or the closely-related “good regulator theorem”.

1. <u>Primary</u> – rules/parameters embodying physiologically determined viral spread features e.g., usual incubation period or duration of infectivity
2. <u>Secondary</u> – rules/parameters determining if/when/where a susceptible person becomes infected or when/where infectious transmission occurs. For example, probability of transmission varies with proximity which we modulate according to two parameters:
  i. ***HazardRadius*** – this parameter functions as a secondary rule to determine transmission risk within a microcosmic spatial reference frame (a single CovidSIMVL “Universe”). Specifically, HazardRadius determines the likelihood of transmission between agents when they are located within specified distances from one another in a given simulation trial step. Protections such as masking would be reflected in lower values on HazardRadius.
  ii. ***MingleFactor*** – this parameter incorporates movements of agents in a delimited “Universe”. Subject to stochastic variation according to a Pareto “random walk” distribution, the MingleFactor parameter, over the course of iterations, determines how many susceptible or infectious agents fall within a critical radius where transmission can take place.
3. <u>Tertiary</u> – rules/parameters that prescribe behaviour for agents with patterned movements between local contexts. **CovidSIMVL *schedules*** describe movements of agents between local Universes as determinants of intra-”universe” interactions which contribute to generation of viral spread dynamics that entail cross-over transmission in the “Multiverse” (array of local contexts).

As demonstrated Moselle & Chang (2020)^26^ higher HazardRadius and MingleFactor values within a finite space (resulting in higher density of agents) produce infection rates summarized by curves associated with higher R_0_ values. These curves need not be as smooth as those typically associated with compartment equation-based models, particularly since discrete-time transitions as entailed by the CovidSIMVL schedule may produce jumps in counts. We stop short of invoking true discontinuity here given the Petri-Net model^27,28^ of continuous discrete event simulations in a finite space.

## METHODS

### CovidSIMVL Trials

CovidSIMVL is an open source, public domain system written in JavaScript and freely available here under the GNU Open License Framework: www.github.com/ecsendmail/MultiverseContagion.

The method employed in this study of transmission chains and superspreader events entails the generation of an array of simulations (trials), keyed to different combinations of CovidSIMVL parameters HazardRadius and MingleFactor. No assumptions about reproduction rate (R_0_) are involved in determining whether viral transmission occurs. Each trial runs through multiple generations until a target value for % infected is achieved. Each simulation is initialized with 100 susceptible agents and one infective agent. The dimensions of the space within which agents interact is held constant for all trials in this study.

- At any point in time, an agent is in one of four states-susceptible, incubating, infective, or inert (e.g., recovered, deceased, or immunized).
- With each generation, agent location will change, and state may change depending on intrinsic agent-based factors (durations associated with incubation and infectivity) or physical location. Neither the rate of transmission nor the length of transmission chains are conditional upon any prior expected values, e.g., an assumed value for R_0_.
- The simulations are run until a target 50% of the initial set of susceptibles are infected, or until the outbreak self-extinguishes i.e., all infectives are inert before the 50% target is met.

In modern browsers such as Chrome, Edge, Safari or Firefox: console log entries are generated at each step of a CovidSIMVL trial, producing a record of every transmission event. Each event is characterized by a sequence number for the iteration/step, a specification of the agents involved in the transmission, and identification of the local spatial context (“Universe”) associated with the event. This information is used to construct (rooted) tree diagrams consisting of chains of transmission generated in the trial.

For reproducibility information within exactness limits imposed by stochastic variation of HazardRadius and MingleFactor please see the technical reports and CovidSIMVL Handbook in the *github* repository mentioned above.

## RESULTS

### Transmission Chains embedded in the CovidSIMVL console.log

As follows we consider traces of transmissions for a synthetic epidemic in the console.log, which provides a window into detailed contagion dynamics. ***Figure 2*** provides a snapshot view of the CovidSIMVL console.log.

**Figure 2.**
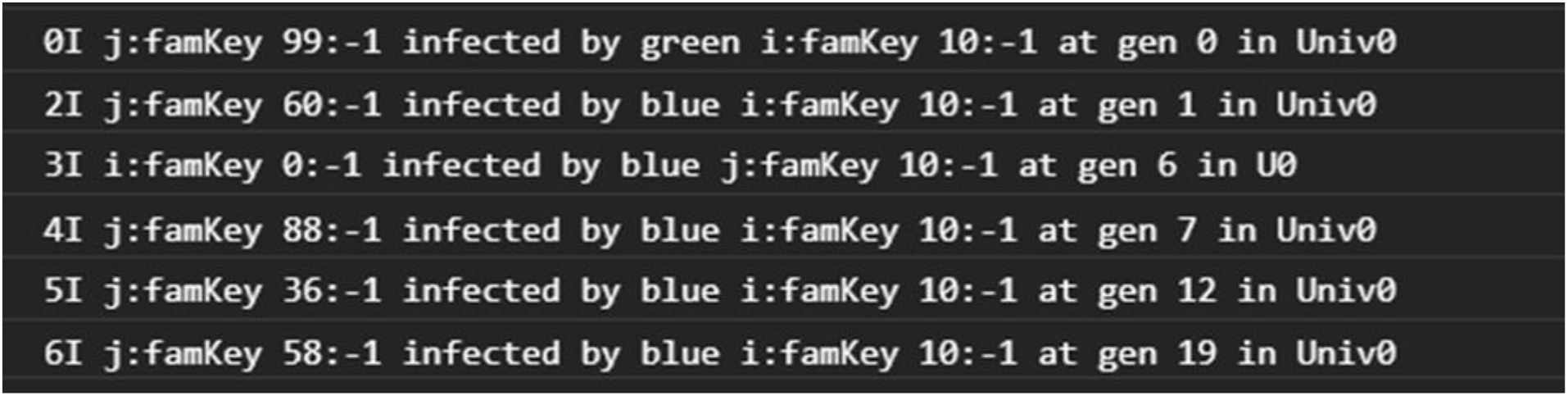
CovidSIMVL console.log

In the following, we use the term “node” to refer to an agent. “Universe” is used to refer to an element of the array of delimited/localized spatial contexts within which one or more agents interact.

The console.log transcript above depicts trial initiation with all events taking place within one Universe. Again, each infection event relates the identities of infected and infective nodes as well as the iteration sequence number for the infection event. For example, node 99 infects node 10 at generation 0 in Universe 0. Then Agent 10 infects 60, 0, 88, 36 and 58 – all in Universe 0.

Likewise from ***Figure 3*** we generate a Transmission Tree in matrix form (***Table 1)*** where the Transmission Trees in that table are sorted by length, not by when the chains were initiated or terminated. In this table, a leaf is the unique terminal node in a chain of transmissions in a rooted tree. For any outbreak where the R_0_ for any agent >1, there will be at least two branches in the tree. The number of distinct chains and the number of leaves (terminal nodes) is the same.

**Figure 3.**
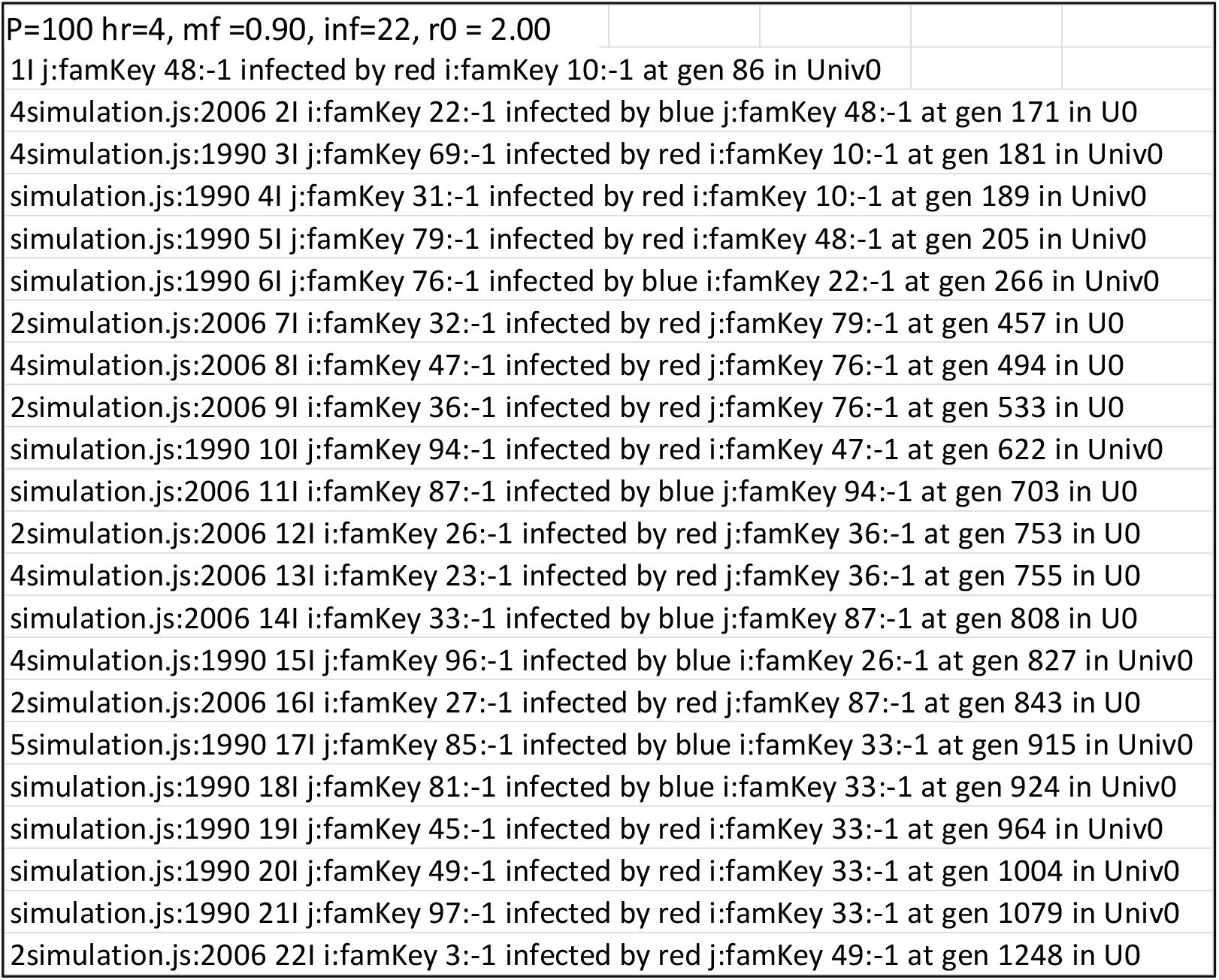
console.log, Population = 100, HazardRadius = 4, MingleFactor= 0.90, First 22 generations

**Table 1.**
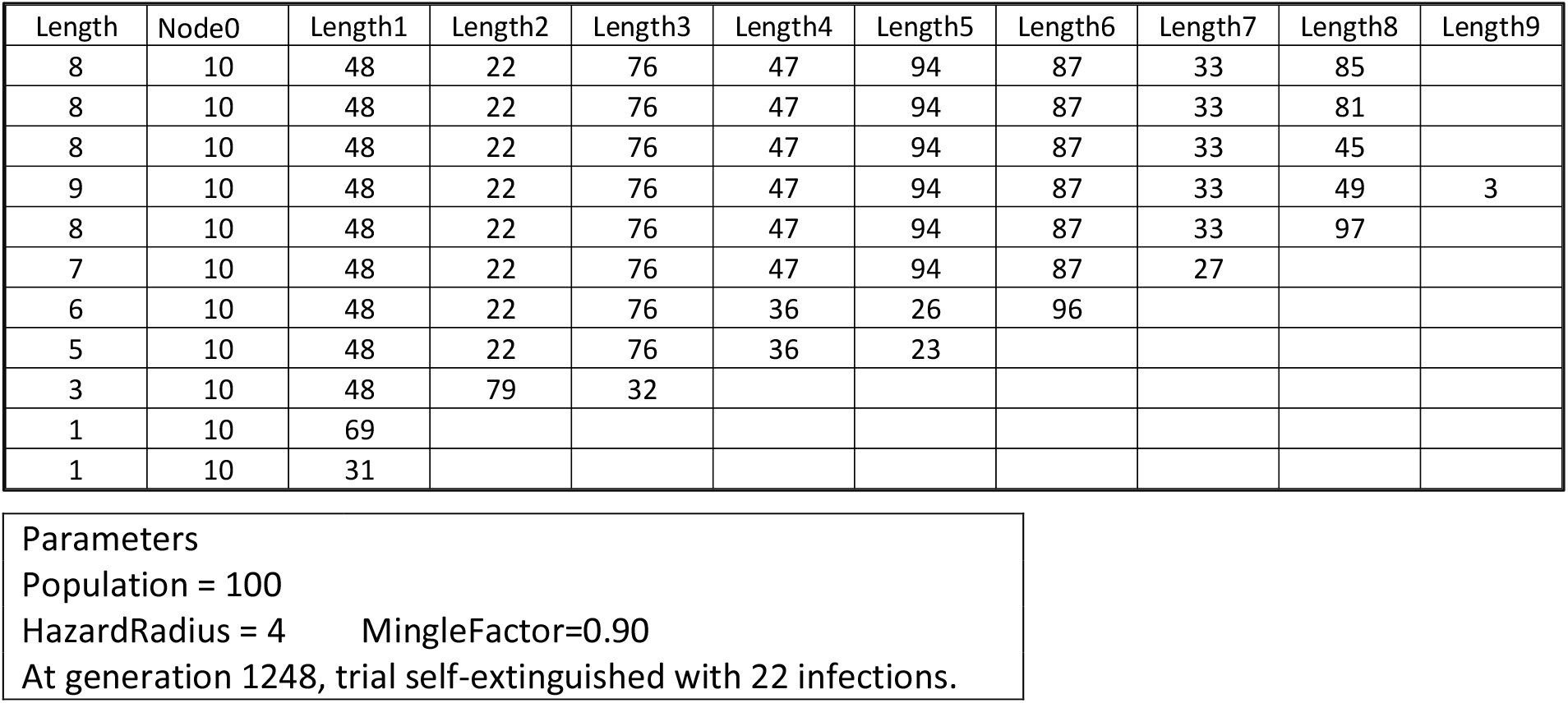
Transmission Tree

| Length | Node0 | Length1 | Length2 | Length3 | Length4 | Length5 | Length6 | Length7 | Length8 | Length9 |
| --- | --- | --- | --- | --- | --- | --- | --- | --- | --- | --- |
| 8 | 10 | 48 | 22 | 76 | 47 | 94 | 87 | 33 | 85 |  |
| 8 | 10 | 48 | 22 | 76 | 47 | 94 | 87 | 33 | 81 |  |
| 8 | 10 | 48 | 22 | 76 | 47 | 94 | 87 | 33 | 45 |  |
| 9 | 10 | 48 | 22 | 76 | 47 | 94 | 87 | 33 | 49 | 3 |
| 8 | 10 | 48 | 22 | 76 | 47 | 94 | 87 | 33 | 97 |  |
| 7 | 10 | 48 | 22 | 76 | 47 | 94 | 87 | 27 |  |  |
| 6 | 10 | 48 | 22 | 76 | 36 | 26 | 96 |  |  |  |
| 5 | 10 | 48 | 22 | 76 | 36 | 23 |  |  |  |  |
| 3 | 10 | 48 | 79 | 32 |  |  |  |  |  |  |
| 1 | 10 | 69 |  |  |  |  |  |  |  |  |
| 1 | 10 | 31 |  |  |  |  |  |  |  |  |

Parameters
Population = 100
HazardRadius = 4 MingleFactor=0.90
At generation 1248, trial self-extinguished with 22 infections.

From the Transmission Tree in ***Table 1***, we can make the following observations:

1. The maximum depth of the tree (maximum path length) is 9.
2. The span of the tree (the number of distinct leaf nodes that have no descendants, i.e., terminal nodes) is 11. This is also the number of distinct chains (or paths to leaves).
3. The unique nodes in this tree is 22. That is the number of people who become infected over a trial. That count does not include infecting the initial infective agent.
4. The trial is set to run until self-extinction or until a threshold value of 50% infected is reached. For the trials in this paper, that would be 50 out of an initial set of 100 susceptibles. For the trial depicted in ***Table 1***, that threshold was reached on iteration 1248. See Moselle & Chang (2020)^29^ for further illustration of the use of a standard threshold value to highlight impacts of systematically varying parameters in the model.
5. An estimate of R_0_ can be generated for each agent that has become inert at the time of termination.
6. Note that in this model R_0_ is not a property set at the initiation of the trial to reflect properties of the virus or a property of a population in a context. It is an emergent property of an individual agent at a specified point in an outbreak, reflecting the consequences of their movements and associated exposure up to that point in the simulation.

### Some assertions on the efficiency of an epidemic

We rate efficiency of an epidemic by the largest number of infections in the shortest duration. In the agent-based model, where the number of transmissions that can take place within a generation is limited by the proximity of agents within a delimited space, the more parallel infective agents that are active, the more quickly a target population is infected.

Thus, balanced trees with less depth (reflecting more infections arising from previous infections) and more breadth (more total leaves) represent efficient epidemic dynamics. For example, a hypothetical balanced tree with 5 descendants from each parent node will be more efficient than a highly unbalanced tree with one long pathway plus many short branches emanating from the same level (generation). At the extreme, a long pathway with no branches and a single terminal node would be the least efficient, as it is a Transmission Tree that happens also to be a single Transmission Chain.

## Transmission Tree generated by different combinations of HazardRadius and MingleFactor

In attempt to clarify and quantify the relationships between the structure of Transmission Trees for parameterized synthetic epidemics, we run a limited number of trials, create potential metrics, and make observations: please see ***Table 2*** for a summary of parameter values and outputs.^30^ The input variables appear in the first 3 columns, indicating the number of susceptibles (Pop) and HazardRadius (HzR) and MingleFactor (MingF) at the point of initialization, prior to any stochastic variation of HazardRadius and MingleFactor parameters. The trials are set to terminate when 50 agents are infected, or when there are no more infectives (i.e., the outbreak self-terminates). EndInf indicates the number of infected agents at trial end.

**Table 2.**
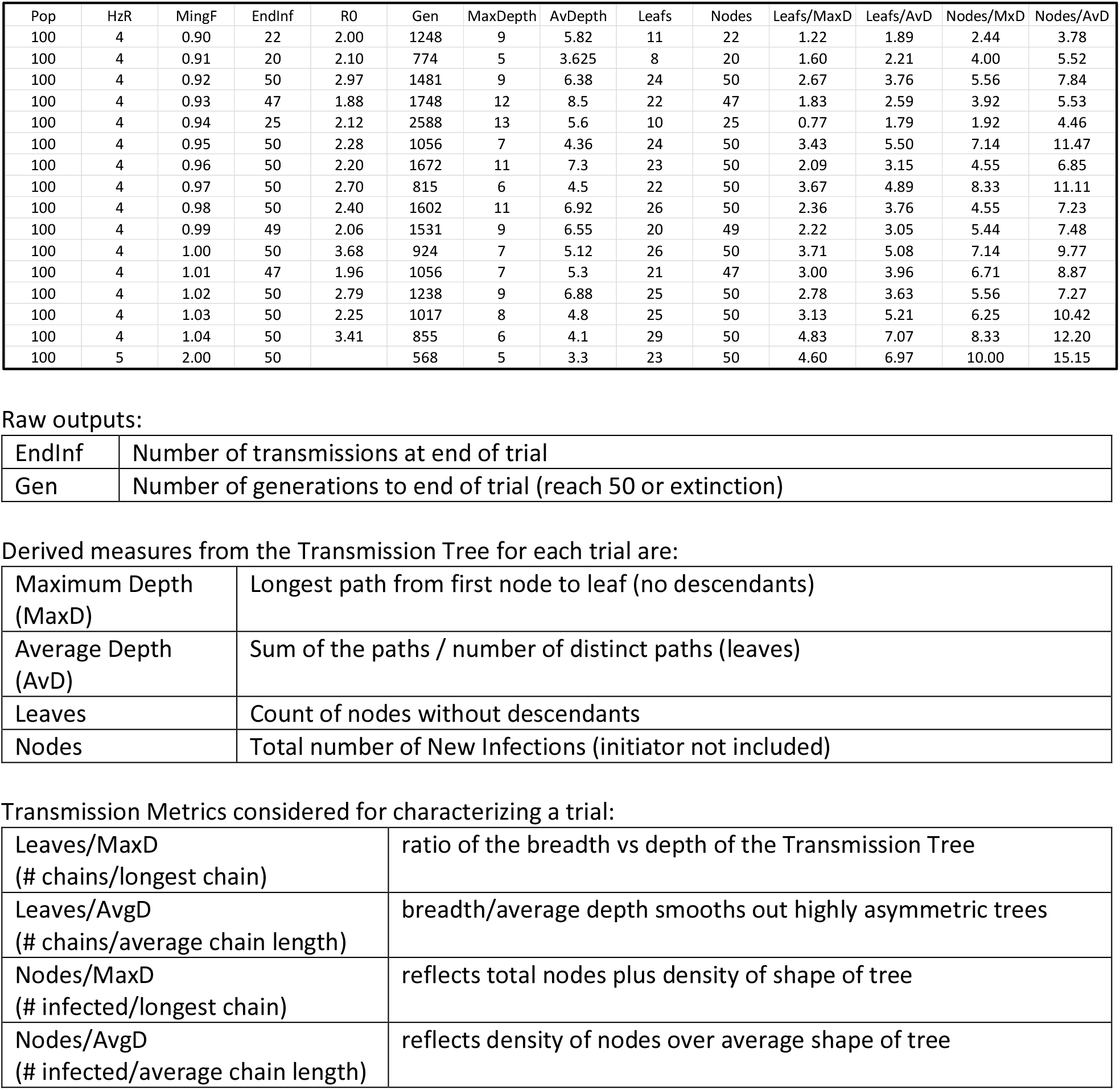
16 Trials – HazardRadius = 4, Varying MingleFactors, Chain Leaves, Nodes and Depth

## Tree Depth as measure of epidemic efficiency

More efficient epidemics spread more quickly (over fewer generations), are embodied in more parallelism at the level of transmission and are reflected topologically in broader, shallower trees. The efficiency is indexed directly by the number of simulated generations (Gen) required to exceed the target threshold of 50%. Here, correlation of Gen with MaxDepth =.94 (P < .001) whereas the correlation is .97 (P < .001) if we consider only 10 out of 16 trials not self-extinguishing before 50% target reached. Along the same lines, the correlation of Gen with AvDepth = .67 (P <. 001), or .95 (P < .001) if we only consider 10 out of 16 trials not self-extinguishing.

## Relationship between agent movement (MingleFactor) and topology of a simulated outbreak

The results in ***Table 3*** suggest that in trials set out in ***Table 2*** there is not a simple relationship between agent movement (MingleFactor) and breadth of tree (i.e., # of Leaves) or length of longest Transmission Chain (Maximum Depth). However, the results are consistent with the assertion that a higher level of agent activity within a delimited space is associated with Transmission Trees that are broader but less deep.

**Table 3.** Correlation of MingleFactor with Transmission Tree topology attributes

| Measures | Correlation | Significance |
| --- | --- | --- |
| MingleFactor with Leaves | .19 | .48 |
| MingleFactor with Maximum Depth (MaxD) | -.40 | .12 |
| MingleFactor with Average Depth (AvgD) | -.43 | .09 |
| Mingle Factor with (Leaves / AvgD) | .57 | .02 |
| MingleFactor with (Leaves / MaxD) | .53 | .03 |

Higher levels of agent movement should produce more efficient spread. Because the trials in ***Table 2*** are stopped at 50% infected, we cannot use these data to verify that MingleFactor does indeed function in this manner in CovidSIMVL. However, the relationship between increasing MingleFactor and increasing R_0_, as reported in Moselle & Chang (2020)^31^ clearly demonstrate the expected relationship.

## Unifying Metrics

For a Transmission Tree, breadth and depth may not cover all the important factors in a synthetic epidemic, which also may include:

1. Number of steps/iterations needed to run the trial to termination – longer trial: less efficient.
2. The number of transmissions before termination – fewer transmissions: less efficient.

In ***Table 2***, the row for MingleFactor 0.97, only 815 generations are needed, compared to >1600 on each side of it, which makes the depth of the tree only 6 instead of 11 for its neighbours.

This epidemic stands out as being more efficient than those on either side. Not seeing any intrinsic reason for 0.97 to be markedly different, we would attribute this result to the stochastic nature of the simulation, which is why the next iteration of CovidSIMVL will enable automated trials and parameter evaluations.

Similarly, examine the trial for MingleFactor 0.94. This level of activity may be expected, based on the trial’s neighbours, to run to 50 new infections but in this trial self-extinguished at 25 infections, using 2588 generations, the largest for all trials.

To incorporate these factors, we add two candidate Unifying Metrics:

1. Unifying Metric **T/E**, defined as:

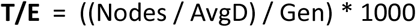

The 1,000 scales to values near 1 to 10. We put the Gen into the denominator so that smaller Generations yield larger T/E efficiencies

2. **Q**, defined as

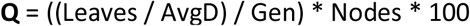

From these definitions, it follows that:

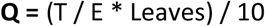

With T/E a measure of efficiency, an epidemic is more efficient if the number of generations required to reach a threshold is smaller, and if the average depth of the Transmission Tree is smaller. Note that “Nodes” corresponds to new infections arising during a trial. Thus, epidemics with more nodes (*ceteris paribus*) are more efficient.

Q modifies the quantity T/E by the number of leaves (which are terminal nodes without descendants). These leaves come from parallel paths: the larger the number of leaves, the more parallelism, hence greater epidemic efficiency.

The two Unifying Metrics’ results are seen in ***Table 4***. In the Green Band, the epidemics are mostly self-extinguishing (EndInf = 25,22,20,47,49) with relatively larger generations except for Gen=1248 and Gen=774 where EndInf are only 22 and 20 respectively. The *Q* values are between 1 and 10.

**Table 4.**
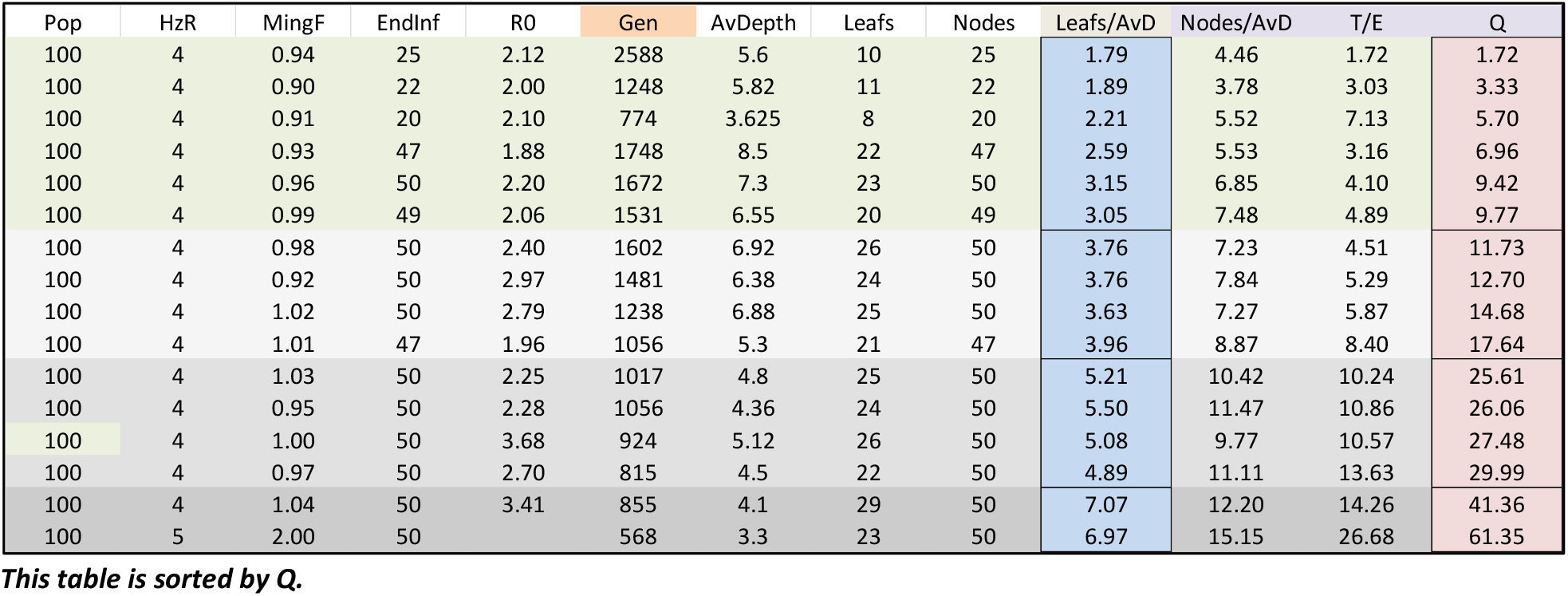
Unifying Metrics

| Pop | HzR | MingF | EndInf | R0 | Gen | AvDepth | Leafs | Nodes | Leafs/AvD | Nodes/AvD | T/E | Q |
| --- | --- | --- | --- | --- | --- | --- | --- | --- | --- | --- | --- | --- |
| 100 | 4 | 0.94 | 25 | 2.12 | 2588 | 5.6 | 10 | 25 | 1.79 | 4.46 | 1.72 | 1.72 |
| 100 | 4 | 0.90 | 22 | 2.00 | 1248 | 5.82 | 11 | 22 | 1.89 | 3.78 | 3.03 | 3.33 |
| 100 | 4 | 0.91 | 20 | 2.10 | 774 | 3.625 | 8 | 20 | 2.21 | 5.52 | 7.13 | 5.70 |
| 100 | 4 | 0.93 | 47 | 1.88 | 1748 | 8.5 | 22 | 47 | 2.59 | 5.53 | 3.16 | 6.96 |
| 100 | 4 | 0.96 | 50 | 2.20 | 1672 | 7.3 | 23 | 50 | 3.15 | 6.85 | 4.10 | 9.42 |
| 100 | 4 | 0.99 | 49 | 2.06 | 1531 | 6.55 | 20 | 49 | 3.05 | 7.48 | 4.89 | 9.77 |
| 100 | 4 | 0.98 | 50 | 2.40 | 1602 | 6.92 | 26 | 50 | 3.76 | 7.23 | 4.51 | 11.73 |
| 100 | 4 | 0.92 | 50 | 2.97 | 1481 | 6.38 | 24 | 50 | 3.76 | 7.84 | 5.29 | 12.70 |
| 100 | 4 | 1.02 | 50 | 2.79 | 1238 | 6.88 | 25 | 50 | 3.63 | 7.27 | 5.87 | 14.68 |
| 100 | 4 | 1.01 | 47 | 1.96 | 1056 | 5.3 | 21 | 47 | 3.96 | 8.87 | 8.40 | 17.64 |
| 100 | 4 | 1.03 | 50 | 2.25 | 1017 | 4.8 | 25 | 50 | 5.21 | 10.42 | 10.24 | 25.61 |
| 100 | 4 | 0.95 | 50 | 2.28 | 1056 | 4.36 | 24 | 50 | 5.50 | 11.47 | 10.86 | 26.06 |
| 100 | 4 | 1.00 | 50 | 3.68 | 924 | 5.12 | 26 | 50 | 5.08 | 9.77 | 10.57 | 27.48 |
| 100 | 4 | 0.97 | 50 | 2.70 | 815 | 4.5 | 22 | 50 | 4.89 | 11.11 | 13.63 | 29.99 |
| 100 | 4 | 1.04 | 50 | 3.41 | 855 | 4.1 | 29 | 50 | 7.07 | 12.20 | 14.26 | 41.36 |
| 100 | 5 | 2.00 | 50 |  | 568 | 3.3 | 23 | 50 | 6.97 | 15.15 | 26.68 | 61.35 |

*This table is sorted by Q.*

In the next band, *Q* values range between 10 and 20. Even though MingleFactor=1.01 does not create 50 new transmissions, extinction is in 1056 generations, with smaller Transmission Tree depth: together these contribute to the epidemic efficiency of *Q*. The stochastic nature produced a rapid epidemic which at 47 generations, ran out of susceptible agents.

The next band of *Q* values, ranging from 20 to 30, are characterized by fewer generations (Gen) and smaller AvgD than the previous two bands. However, *Q* is not statistically related to MingleFactor (correlations of MingleFactor with Q = −.26, P = .34) so *Q* is driven by actual epidemic progress, rather than the parameters, which set the framework but not the unfolding in a stochastic universe.

The final band with *Q* values above 40, clearly have fewer generations, and shallower Transmission Tree depths, reflecting overall efficiency.

These observations are for single trials at various parameter settings. The stochastic nature of CovidSIMVL implies that more observations at these settings would permit confidence level estimates.

## CONSIDERATIONS for R_0_

The approach we have taken for R_0_ is to record the number of susceptibles each agent successfully infects. However, when we terminate at newInf = 50, there may be a significant number of agents that have not completed their life cycle. We know that we have counts for those which are inert, and we can average over those to provide a value of R_0_.

These are of course early infectives, as susceptibles are never inert. However, the random nature of the simulation does not guarantee that all populations to any point have the same number of infectives.

By examining the Transmission Trees, we see many that do not have a balanced structure. If the distribution of infections are random, any particular R_0_ at newInf = 50 or at extinction is not necessarily reflective of the average over a large number of trials with a given parameter setting of Population, HazardRadius and MingleFactor.

### Rooted tree structure and shared paths

Epidemics with more serial than parallel executions would share more paths that have common ancestors, until a branching point is reached. This characteristic may be of use in describing the epidemic dynamics, if we can capture the relationships.

With lower degrees of parallelism, there would be more common paths, and longer ones, such as in the Transmission Tree for HazardRadius (HzR) = 4 and MingleFactor (mF) = 0.94, with a *Q* of 1.72 – see ***Figure 4***, below.

**Figure 4.**
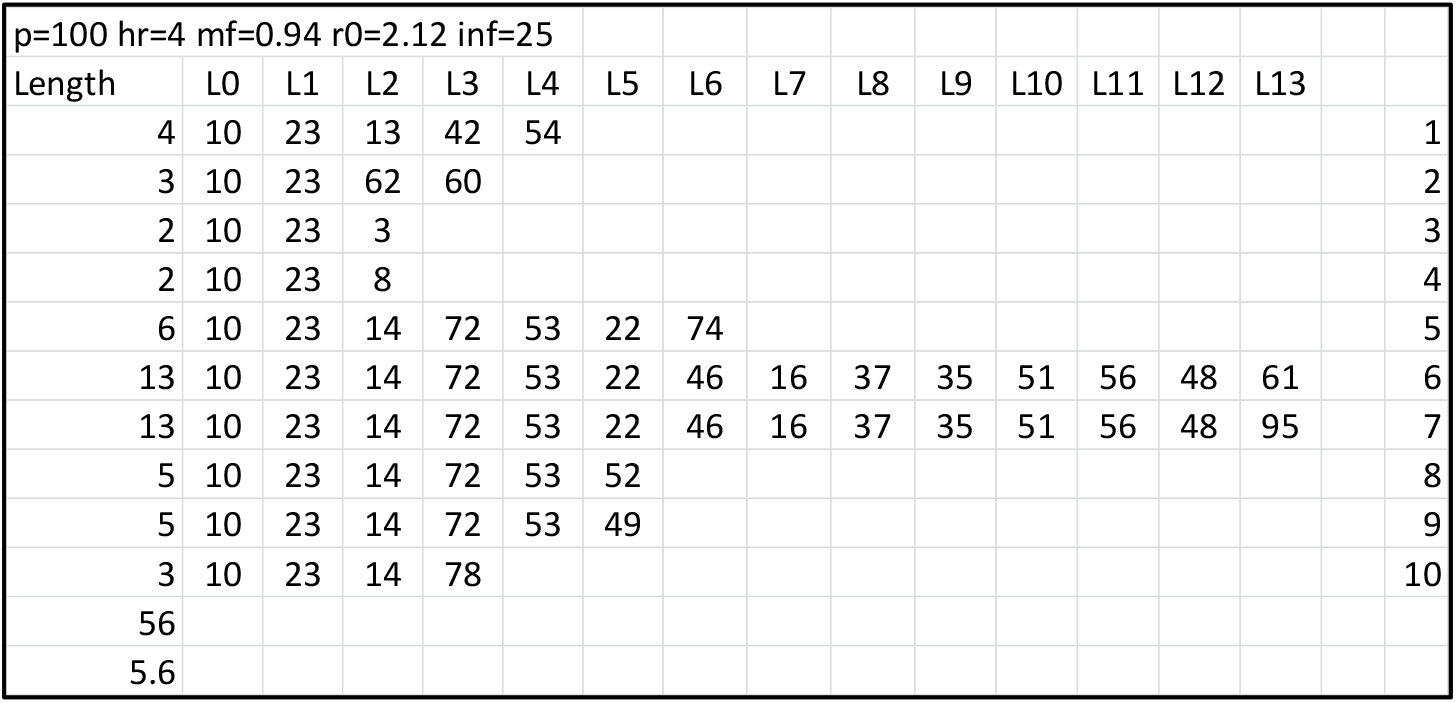
Transmission Tree, HazardRadius = 4, MingleFactor = 0.94, Q = 1.72

On the other hand for HzR = 5 and mF = 2.0 we have high Q = 61 (very efficient epidemic) with many shorter paths. Longer common paths are signs of a slower epidemic. See ***Figure 5***.

**Figure 5.**
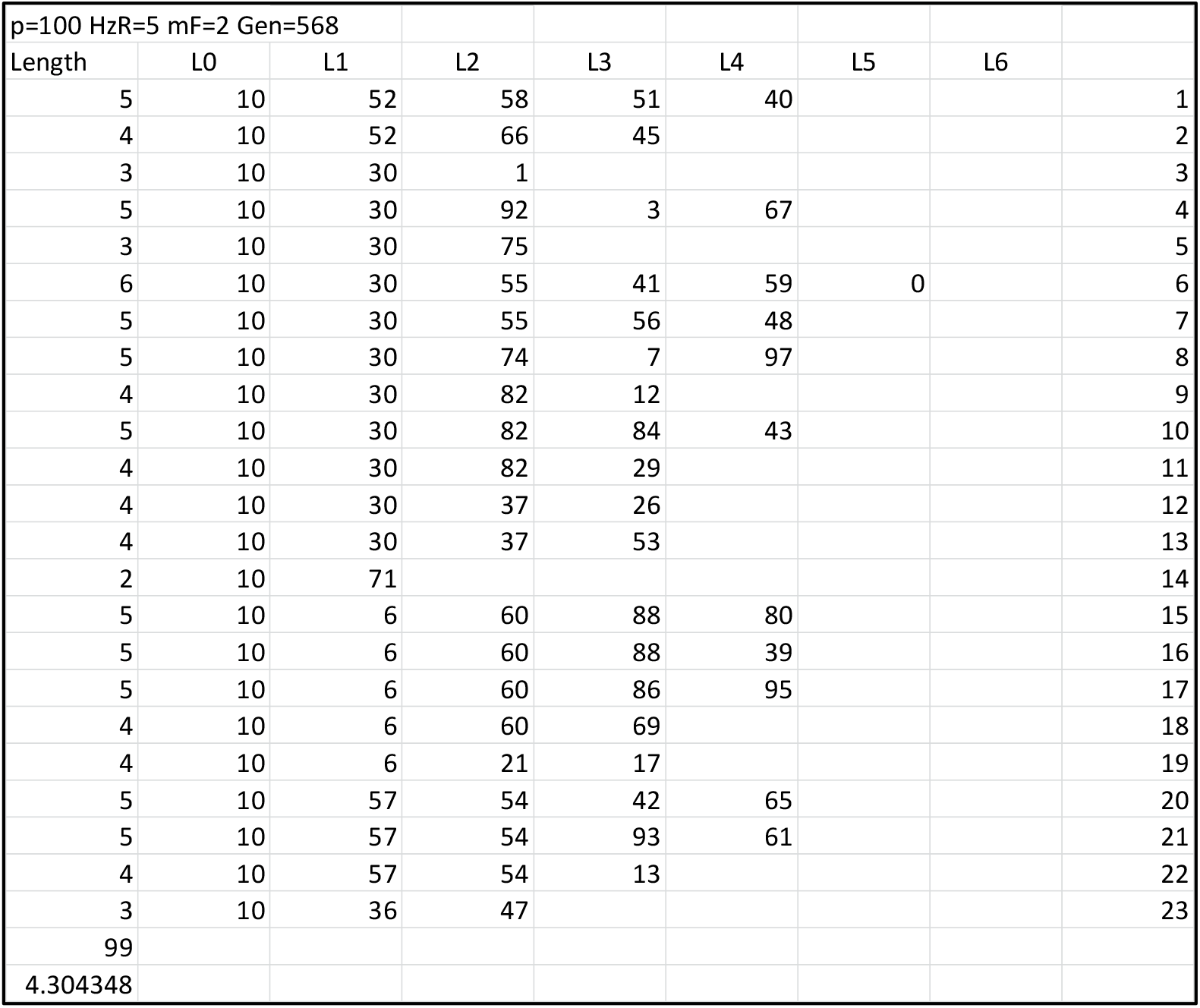
Transmission Tree, HazardRadius = 5, MingleFactor = 2.0, Q = 61

### Degree of Infectivity of Agents

There are two ways to look at the role individual agents (or persons) can have in an epidemic. One is ro count the persons infected as a consequence of the individual being infective. This corresponds to tallying the size of the sub-tree of descendants of the individual in the Transmission Tree. We term this the *Spread Count* of an infective agent.

Another is to count an agent’s *direct* descendants, rather than all descendants of a given agent. We refer to this as *Case Count*, which corresponds to the reproduction number (R_0_) for an individual agent *at a given stage* (generation) in an outbreak.

To illustrate these two concepts, we compare two Transmission Trees, taken from the ***Supplement***, for HzR = 4, MingleFactor = 0.90 and MingleFactor = 1.04. The first (mF = 0.90) is a trial self-extinguishing after 22 new infections and 1,248 generations, with R_0_ of 2.00. The second (mF = 1.04) completes 50 new infections in 855 generations with a terminating average R_0_ of 3.41.

The two Transmission Trees taken from the ***Supplement*** are depicted in ***Figure 6. Table 5*** summarizes differences between the two Transmission Trees.

**Figure 6.**
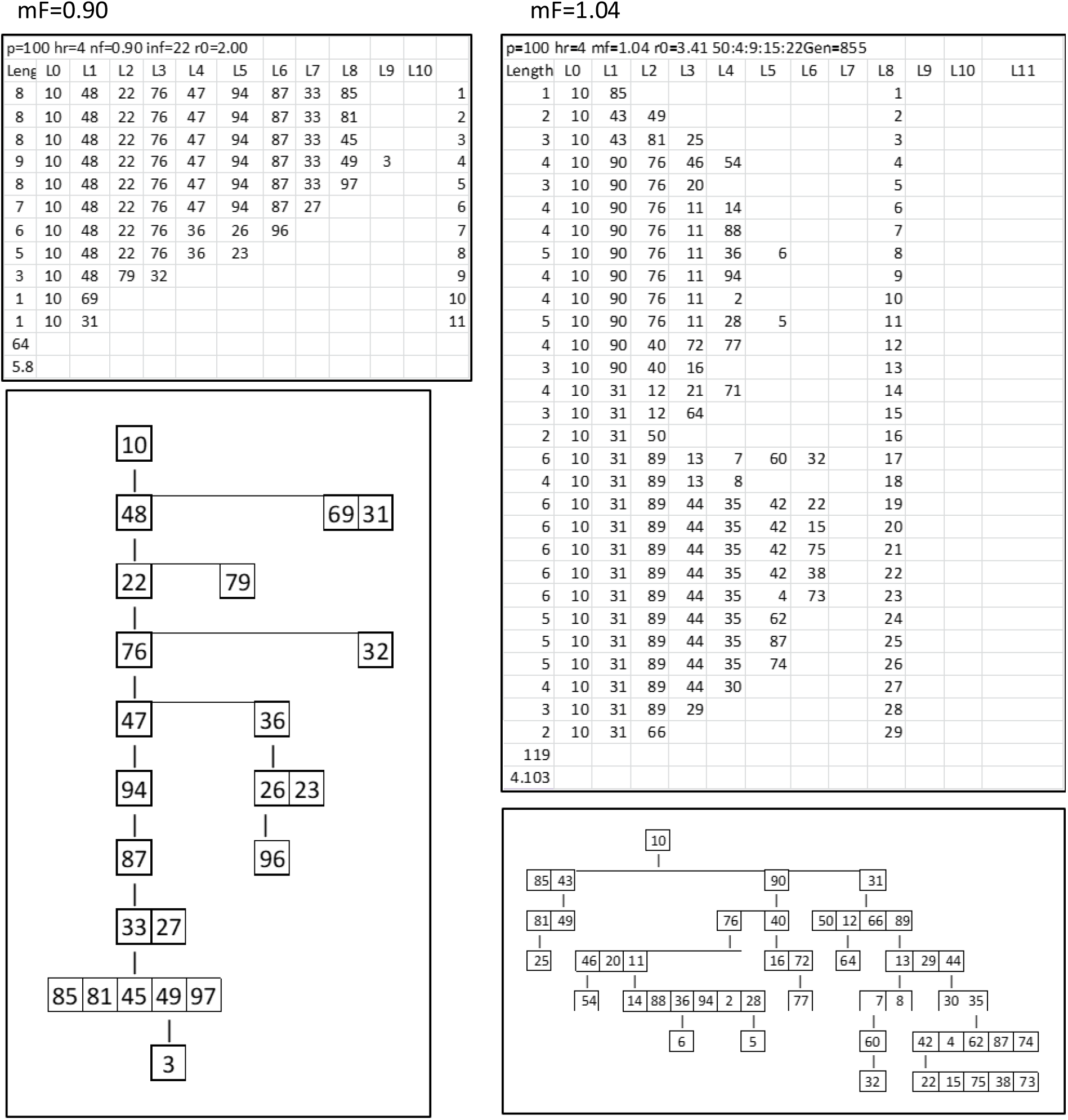
Two Transmission Trees for HazardRadius = 4 and mF = 0.90 or mF = 1.04.

**Table 5.**
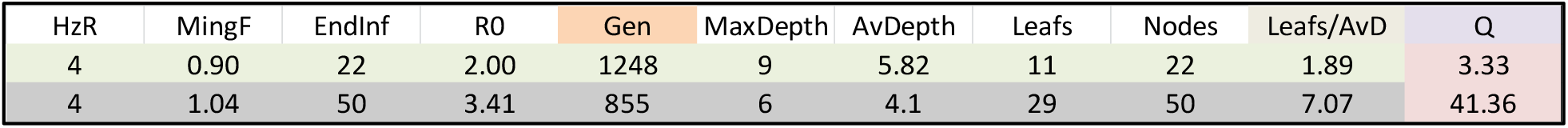
Summary statistics for Transmission Trees generated by mF = 0.90 and mF = 1.04

| HzR | MingF | EndInf | R0 | Gen | MaxDepth | AvDepth | Leafs | Nodes | Leafs/AvD | Q |
| --- | --- | --- | --- | --- | --- | --- | --- | --- | --- | --- |
| 4 | 0.90 | 22 | 2.00 | 1248 | 9 | 5.82 | 11 | 22 | 1.89 | 3.33 |
| 4 | 1.04 | 50 | 3.41 | 855 | 6 | 4.1 | 29 | 50 | 7.07 | 41.36 |

The first case (the mF = 0.90 trial) has *Q* = 3.33, extreme inefficiency, where the outbreak self-extinguishes after 1,248 Generations due to stochasticity (reflected in variation in case counts) plus less physical movement within a finite space occupied at the time of initiation by 100 agents. The second case (mF = 1.04) has *Q* factor 41.36, representing a very efficient outbreak. This second case hits the trial termination threshold of 50% after 852 Generations.

We compare Spread Counts and Case Counts for the two trees, as depicted in ***Figure 7***.

**Figure 7.**
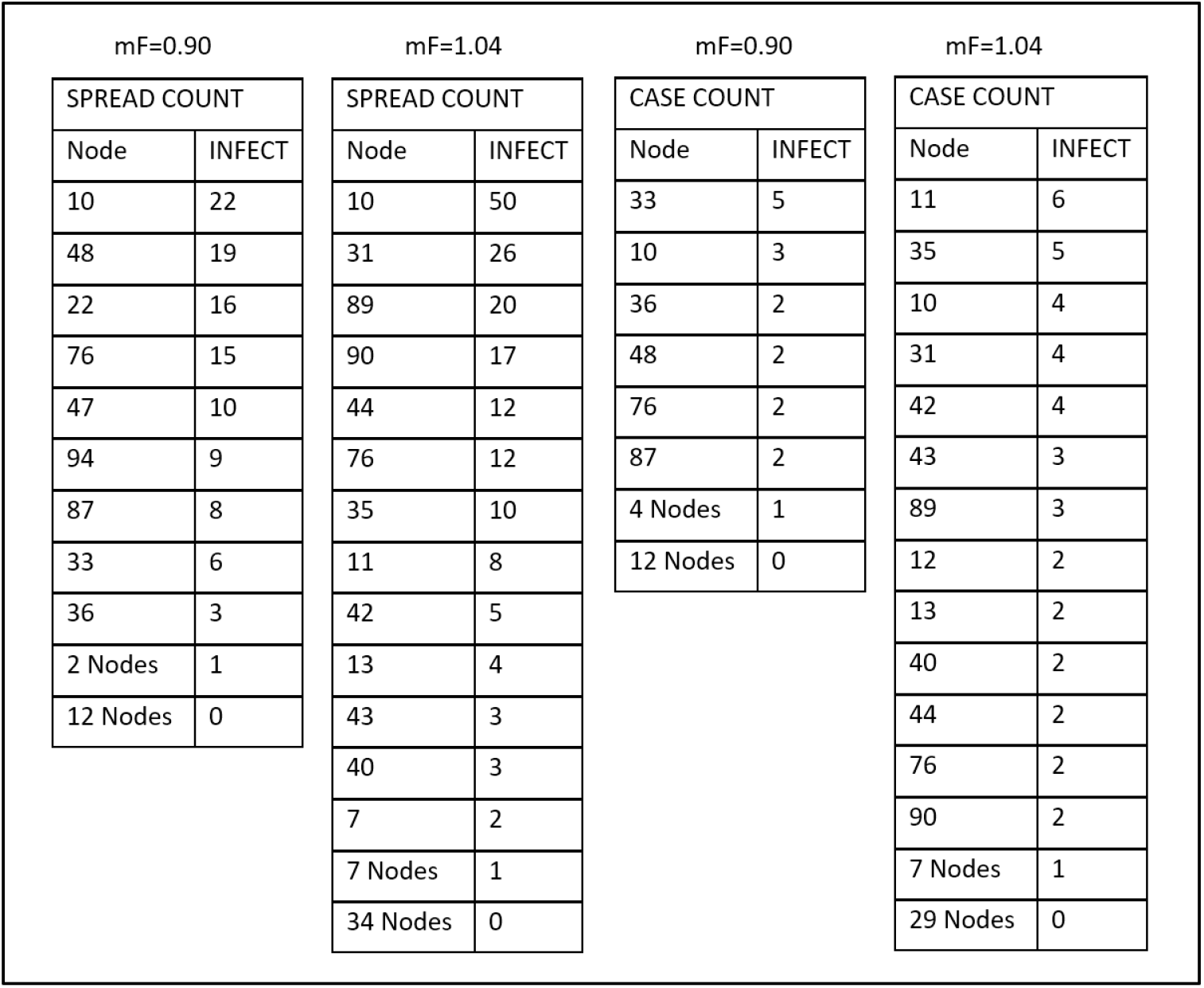
Spread Counts and Case Counts for mF = 0.90 and mF = 1.04 trials

The mF = 1.04 trial has a larger leaf to AvgD ratio at 7.07, compared to 1.89, which shows a higher degree of parallel activity. The Transmission Tree for this trial is broad and shallow. The nodes higher up have larger sub-trees, so it will take fewer nodes to account for any given proportion of infected.

Note that for uniform spread we might expect Spread Count for each node at a particular generation in the simulation (and hence, at the same tree level) to have the same number of offspring. However, given the stochasticity built into each transmission event for CovidSIMVL, at each generation we have agents who are infected at the same generation in a trial associated with different numbers of offspring.

The Transmission Tree for the mF = 0.90 trial is tall and skinny, with only one node having more than three descendants.

Plotting the Spread Counts for the mF = 0.90 trial (***Figure 8)*** we see that the Spread Counts show a steady roughly linear decline, where larger Spread values are associated largely with the timing of an agent becoming infected.

**Figure 8.**
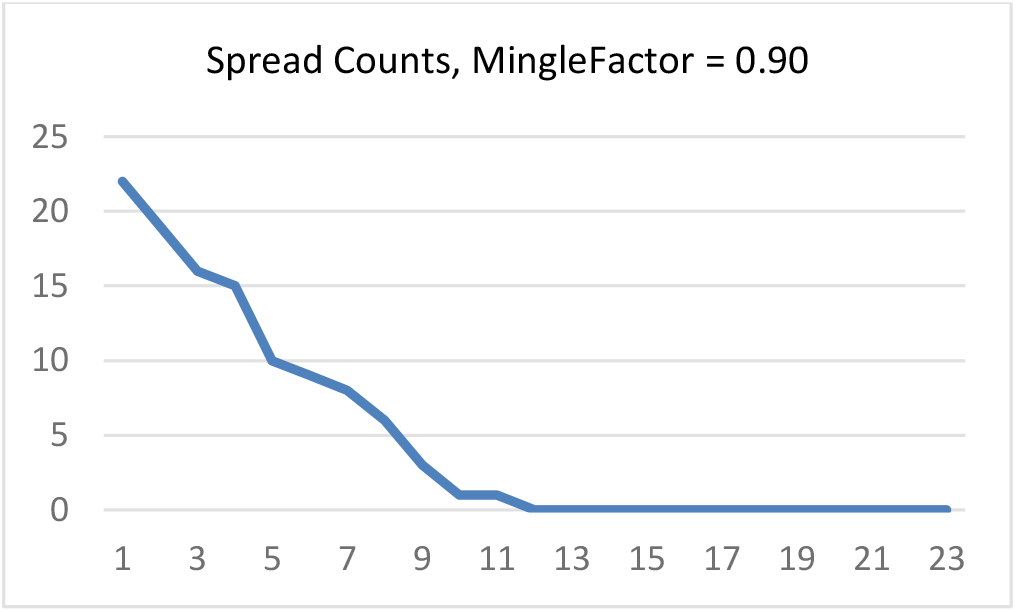
Spread Counts, MingleFactor = 0.90

By contrast, see plotted Spread Counts for mF = 1.04 in ***Figure 9***. We find a curve resembling expected negative binomial offspring distributions arising from branching processes in over-dispersed count data (i.e., low binomial dispersion parameter *k*^32^). Plotted Spread Count values transformed with the natural logarithm in ***Figure 9*** show expected linearity.

**Figure 9.**
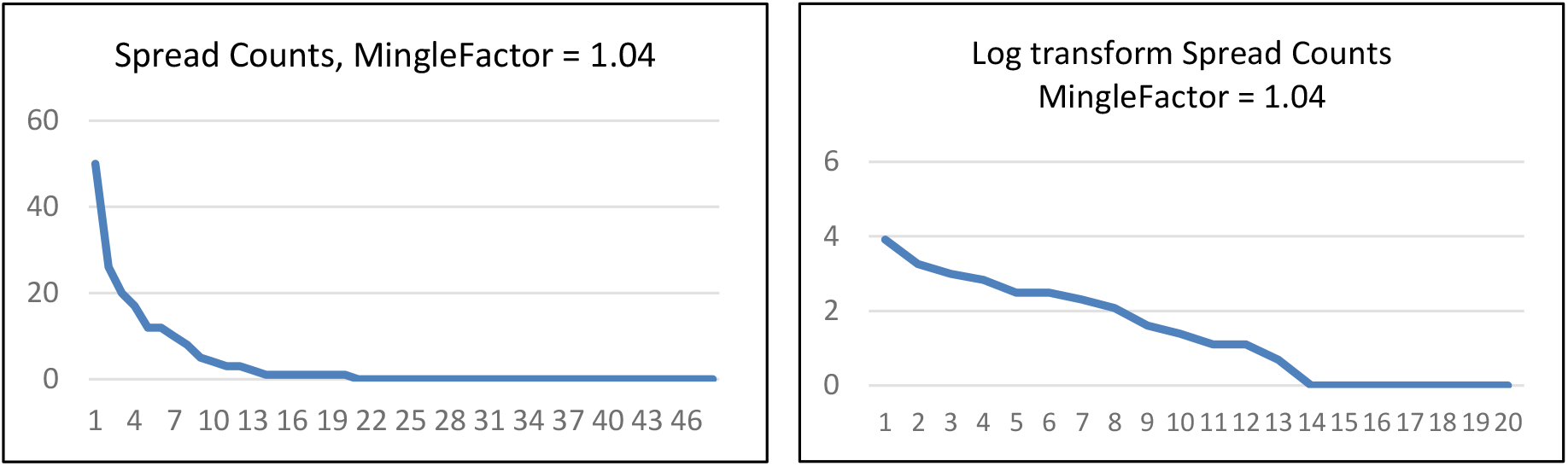
Spread Counts, Natural Logarithm of Spread Counts, MingleFactor = 1.04

Consistent with the “80-20” rule associated with these types of distributions^33^ we find a large proportion of infections associated with a small fraction of the total set of agents. We interpret this to be a consequence of a higher degree of movement by agents (mF = 1.04 vs 0.90) resulting in more rapid spread relative to infective duration. This results in a critical threshold for sustained infection being crossed^34^ so the outbreak does not self-extinguish, resulting in a larger net total number of cases being infected--50 infections over 852 generations *vs* 22 infections over 1,248 generations for the mF = 0.90 outbreak.

For uniform spread we might expect equal Spread Counts for each person (each person spreads to the same number of descendant infected) but with the stochastic tree of primary and secondary and tertiary spread, inevitably some agents infect more descendants than others. When we sort the agents by descendant count we get some agents with many descendants and some with fewer.

Regarding Case Count the mF=1.04 trial, stopped at 50% infection of original susceptibles has 13 agents primarily infecting 2 or more agents. For mF = 0.90 trial, self-extinguishing with 22 agents infected, there are 6 agents infecting 2 or more agents.

Both trials use the same HazardRadius value, embodying stochastic variation in viral load and duration of infectivity. Variation in Case Count in-trial reflects stochastic variation in the HazardRadius factor.

Mean Case Count differences reflect differences in the R_0_ values, which are 2.00 for the mF = 0.90 trial *vs* 3.14 for the mF = 1.04 trial (see ***Table 5***, above). Note that despite R_0_ of 2.00, the mF = 0.90 trial self-extinguishes after 22 of 100 original susceptibles are infected. For mF = 1.04 trial, transmissions continue, with the trial halted when threshold of 50% infected is reached.

These differences in R_0_ values and Case Counts for mF = 0.90 trial *vs* the mF = 1.04 trial illustrates impact of the mF (MingleFactor parameter) both on the efficiency and sustainment of transmission, and on number of primary infections associated with any given infective agent. This points to contribution of physical movement of agents in space to the Case Count differences.

### Comment on Superspreaders

If by “superspreader” we mean agent with high degree of primary or *direct* infection, relative to the average or typical infective, the mF = 0.90 trial and the mF = 1.04 trial do produce some variation in number of direct infections, with statistical outliers as shown in ***Figure 10***. For direct infections depicted in the figure, the shape of the curves is similar, reflecting the fact that the HazardRadius is held constant across the two trials, though it is subject to stochastic variation.

**Figure 10.**
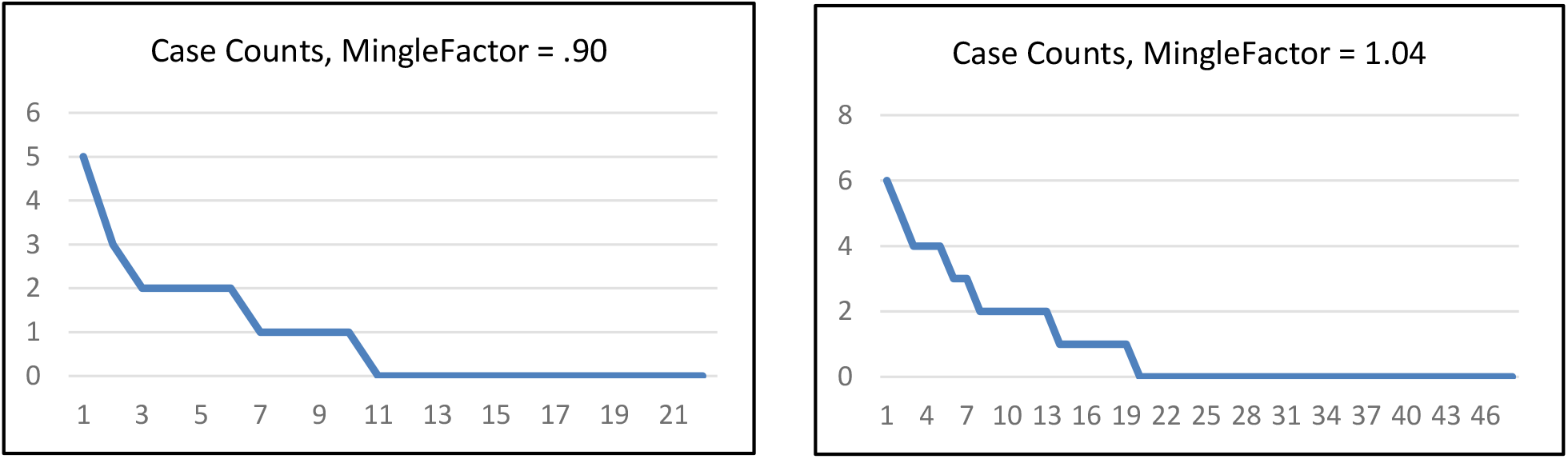
Case Counts, mF = 0.90 and mf = 1.04 trials.

HazardRadius may be understood as a property “owned” by agents, producing a main effect independent of movement of agents within a space, though there are also interactions between HazardRadius and the effect generated by movement. To accentuate the superspreader dynamic in a simulation, the MingleFactor would also need to be set at a level ensuring a requisite portion of space(s) covered by infectives and shared with susceptibles is large enough, and the space is covered sufficiently rapidly that the infection does not self-extinguish before any given infective has transmitted to a large number of susceptibles occupying that space.

On the other hand, if “superspreading” counts all descendants infected by primary and secondary (and so on) infections along all subsequent infection chains, then the stochastic nature of the agent based model guarantees significant asymmetry, if for no other reason than that spread unfolds over the course of generations with values of parameters stochastically varying.

These may not physiologically be “superspreader” events but statistical artifacts of stochastic and probabilistic transmissions based on moving contacts, and on spread unfolding over the course of generations. We can have “superspreader” events without superspreading individuals.

### Comment on Superspreader Events

There are three ways that the term “superspreader” finds common use. They are:

1. A highly infective person creating many ***primary*** infections
2. An infection source for (eventually) large numbers cases who may be “held responsible”
3. An event in which “unexpectedly many” new cases arise.

With regard to the first use, the current literature sometimes references viral load as a possible mechanism, which may also relate to duration of infectivity.^35,36,37^ This parameter in CovidSIMVL reflecting viral load is subject to stochastic variation over generations within trials. This is reflected in variability in the Case Counts metric in this paper.

Regarding the second use, the Spread Count metric provides a direct measure.

For the third use, where the number of cases associated with an event or a context is statistically elevated relative to some modal value, CovidSIMVL can be configured to produce such simulations by setting values on HazardRadius and MingleFactor such that the virus does not self-extinguish early, and by increasing the number of infective agents used to prime the simulation. Note that in the case of the material above, the simulations are primed with just one infected agent.

In the ***Figure 11***, we have results from the first iteration (Gen) of a CovidSIMVL trial starting with 100 susceptible plus 5 infective persons, a Hazard Radius of 20 and MingleFactor = 10.

**Figure 11.**
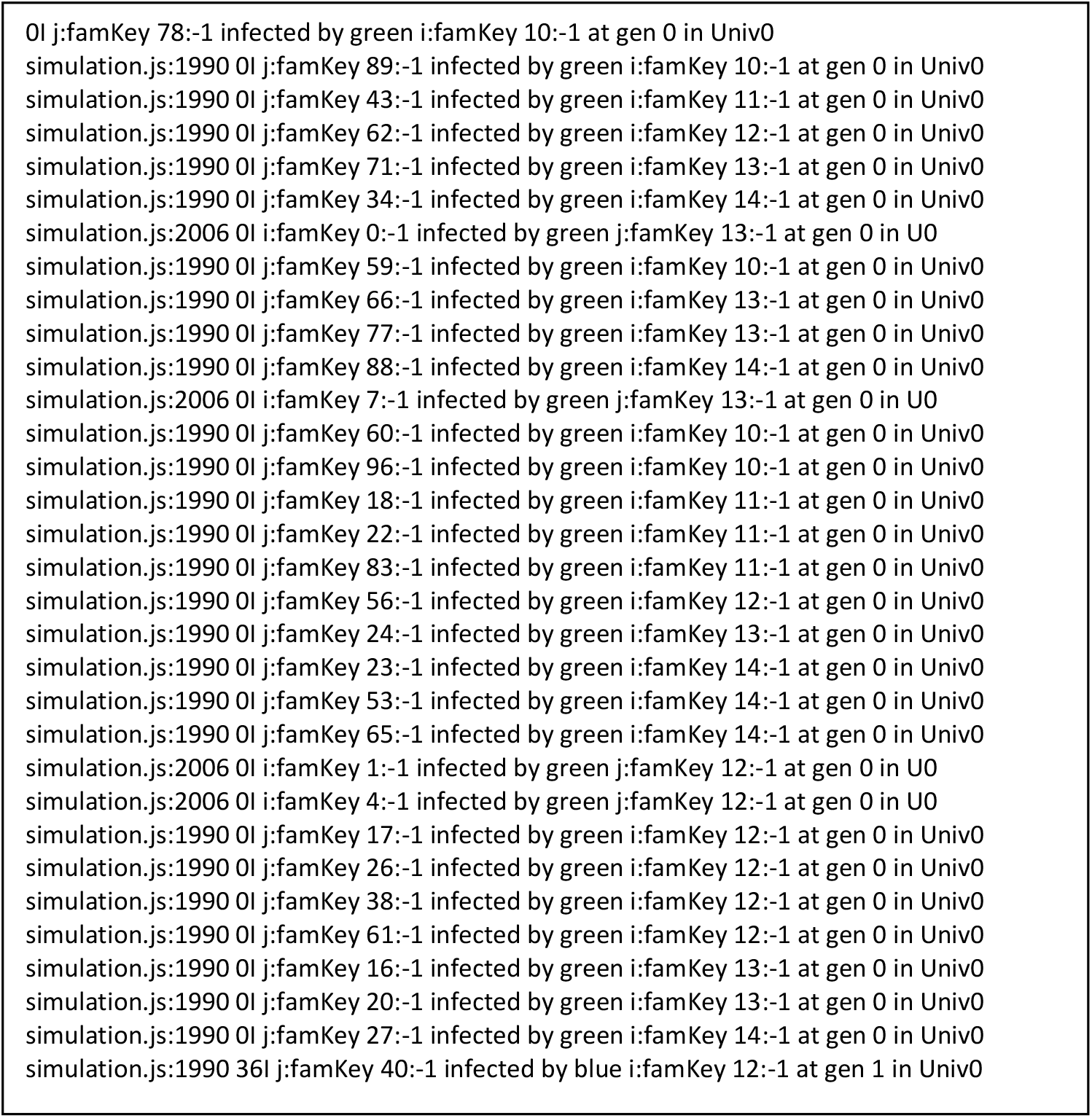
First Iteration (Gen) transmissions, HazardRadius = 20, MingleFactor = 10, 5 Infectives.

This figure shows 31 persons infected by five active cases in the first hour, distributed among agents 10 to 14 with Case Counts = 5, 4, 8, 8, 6 for these five agents. This is a simple demonstration of the power of density and mingling in the contagion spread of SARS-CoV-2 when agents are infective and moving in such a way that the mechanisms of transmission cover the distance between infective and susceptible.

For this same trial, ***Figure 12*** depicts transmission after 2 generations. In this graphic depiction of infection spread generated by CovidSIMVL BLUE = initial infective, YELLOW=infected and GREEN = susceptible.

**Figure 12.**
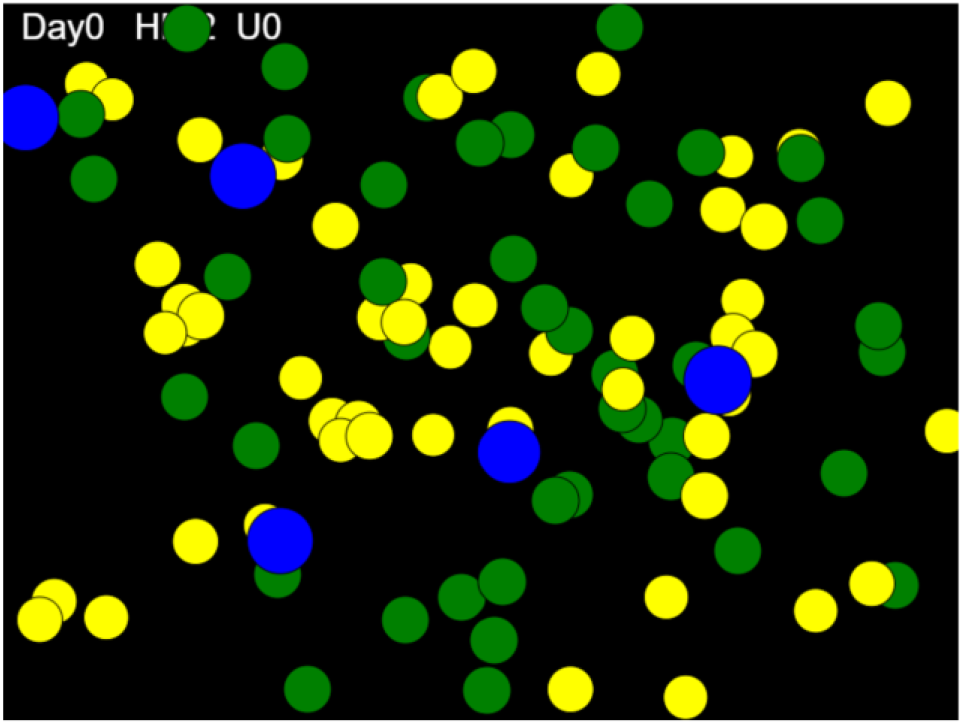
Second Iteration (Gen) transmissions, HazardRadius = 20, MingleFactor = 10, 5 Infectives.

## CONCLUSION – Potential Applications of the Tree Characterization of Viral Transmission

It is intuitively clear that an epidemic spreading rapidly through a population has many infective agents, that chains of transmission are short (if we can trace them) and that leaves (terminal points of Transmission Chains) at any one time are plentiful compared to the relative length of the chains in slower-growing epidemics.

Looking at contact tracing as sample survey of chains of transmission, if these chains are examined in aggregate form reconstructing the Transmission Tree or Trees, we could estimate Generation, Depth (path length), Leaves and New Infections, and thus calculate measure of efficiency (*Q*) as we defined.

Often it is somewhat difficult to know with the variations in daily counts and positivity whether the dynamic at work at a particular time will increase likelihood of transmission rate rising, falling or remaining similar. By looking at the topology of Transmission Trees and the distribution of trees of different spreads, it may be possible to infer characteristics of the dynamic in a way that is not based purely on infection counts or positivity rates. This may be a preferred option in early stages of an outbreak when rates are low and proportionately quite variable from one day to another, and when confidence intervals around equations fit to those data points may be quite large.

Contact tracing may be seen as forward or future-facing activity. Given a person who tests positive, we want to know who they have contacted during a period of presumed infectivity, prior to receipt of test positive results, so we can know who needs testing, monitoring or possibly treatment. The objective is to identify all persons that the Index Case might infect.

Facing backwards the concern is how a test-positive case is positioned in transmission chain. A “side-effect” of such retrospective investigation is identification of persons contacted by the Index Case during an infective period prior to identification (and presumably isolation).

However, an important objective of this backward-facing approach to contact tracing would be to build out a picture of distributions of chains of transmission. This investigation would need to go as far back as the length of typical and atypical chains dictated. It would need to go as far afield as necessary to generate a stable depiction of typical and atypical spread of Transmission Trees.

This information could then be employed to generate measures related to transmission dynamics – velocity or efficiency, and topology of transmission or distribution of cases within some meaningfully demarcated geographical entity or some other factor (e.g., a workplace or co-location in a permanent or transient congregate living situation) networking people together. Technology may in principle be employed to link co-location and movement of people to test results, providing an alerting mechanism for infections in early stages of escalation, when equation-based models may be less well-suited to describing and predicting.^38,39^

## Data Availability

The simulating modeling tool is available on a github site. The location for that site appears in the paper. All tables of results that are summarized in the document are included in a supplementary document to be submitted to medRxiv

https://github.com/ecsendmail/MultiverseContagion

