## Supplemental material for "CovidSIMVL --Transmission Trees, Superspreaders and Contact Tracing in Agent Based Models of Covid-19"

**Affiliations:**
[a]Consultant
[b]Island Health (Vancouver Island Health Authority, BC, Canada); University of Victoria, Departments of Health Information Science, Psychology
[c]Province of British Columbia

*

**Running Title**: CovidSIMVL Transmission Trees, Superspreaders and Contact Tracing
**Keywords:** SARS-Cov-2, COVID-19, agent-based model, superspreader, network, contact tracing

This document contains transmission trees for trials discussed in Chang, Moselle & Richardson (2020*)* *CovidSIMVL – Transmission Trees, Superspreaders and Contact Tracing in Agent Based Models of Covid-19*, medRxiv.

### Transmission Trees

| p=100 hr=4 nf=0.90 inf=22 r0=2.00 | | | | | | | | | | | | |
|---|---|---|---|---|---|---|---|---|---|---|---|---|
| Leng | L0 | L1 | L2 | L3 | L4 | L5 | L6 | L7 | L8 | L9 | L10 | |
| 8 | 10 | 48 | 22 | 76 | 47 | 94 | 87 | 33 | 85 | | | 1 |
| 8 | 10 | 48 | 22 | 76 | 47 | 94 | 87 | 33 | 81 | | | 2 |
| 8 | 10 | 48 | 22 | 76 | 47 | 94 | 87 | 33 | 45 | | | 3 |
| 9 | 10 | 48 | 22 | 76 | 47 | 94 | 87 | 33 | 49 | 3 | | 4 |
| 8 | 10 | 48 | 22 | 76 | 47 | 94 | 87 | 33 | 97 | | | 5 |
| 7 | 10 | 48 | 22 | 76 | 47 | 94 | 87 | 27 | | | | 6 |
| 6 | 10 | 48 | 22 | 76 | 36 | 26 | 96 | | | | | 7 |
| 5 | 10 | 48 | 22 | 76 | 36 | 23 | | | | | | 8 |
| 3 | 10 | 48 | 79 | 32 | | | | | | | | 9 |
| 1 | 10 | 69 | | | | | | | | | | 10 |
| 1 | 10 | 31 | | | | | | | | | | 11 |
| 64 | | | | | | | | | | | | |
| 5.8 | | | | | | | | | | | | |

Generations = 1248

| p=100 hr=4 nf=0.91 inf=20 r0=2.10 | | | | | | | | |
|---|---|---|---|---|---|---|---|---|
| Length | L0 | L1 | L2 | L3 | L4 | L5 | L6 | |
| 4 | 10 | 81 | 65 | 56 | 24 | | | 1 |
| 5 | 10 | 81 | 30 | 69 | 7 | 45 | | 2 |
| 5 | 10 | 81 | 30 | 69 | 57 | 12 | | 3 |
| 2 | 10 | 81 | 97 | | | | | 4 |
| 4 | 10 | 37 | 23 | 94 | 95 | | | 5 |
| 3 | 10 | 37 | 35 | 18 | | | | 6 |
| 3 | 10 | 37 | 35 | 73 | | | | 7 |
| 3 | 10 | 37 | 35 | 55 | 29 | | | 8 |
| 29 | | | | | | | | |
| 3.625 | | | | | | | | |

Generations = 774

| | | | | | | | | | | | | |
|---|---|---|---|---|---|---|---|---|---|---|---|---|
| p=100 hr=4 nf=0.92 r0=2.97 inf=50 | | | | | | | | | | | | |
| Length | L0 | L1 | L2 | L3 | L4 | L5 | L6 | L7 | L8 | L9 | | |
| 1 | 10 | 29 | | | | | | | | | | 1 |
| 3 | 10 | 96 | 48 | 2 | | | | | | | | 2 |
| 2 | 10 | 96 | 68 | | | | | | | | | 3 |
| 3 | 10 | 93 | 25 | 37 | | | | | | | | 4 |
| 6 | 10 | 93 | 25 | 20 | 11 | 84 | 69 | | | | | 5 |
| 6 | 10 | 93 | 25 | 20 | 11 | 84 | 94 | | | | | 6 |
| 8 | 10 | 93 | 25 | 20 | 11 | 1 | 15 | 90 | 28 | | | 7 |
| 8 | 10 | 93 | 25 | 20 | 11 | 1 | 15 | 90 | 5 | | | 8 |
| 9 | 10 | 93 | 25 | 20 | 11 | 1 | 15 | 90 | 30 | 59 | | 9 |
| 7 | 10 | 93 | 25 | 20 | 11 | 1 | 15 | 14 | | | | 10 |
| 8 | 10 | 93 | 25 | 78 | 62 | 33 | 66 | 82 | 65 | | | 11 |
| 8 | 10 | 93 | 25 | 78 | 62 | 33 | 66 | 82 | 67 | | | 12 |
| 7 | 10 | 93 | 25 | 78 | 62 | 33 | 79 | 7 | | | | 13 |
| 7 | 10 | 93 | 25 | 78 | 62 | 33 | 53 | 74 | | | | 14 |
| 9 | 10 | 93 | 25 | 78 | 62 | 33 | 24 | 61 | 31 | 57 | | 15 |
| 9 | 10 | 93 | 25 | 78 | 62 | 33 | 24 | 61 | 31 | 71 | | 16 |
| 7 | 10 | 93 | 25 | 78 | 62 | 33 | 24 | 21 | | | | 17 |
| 7 | 10 | 93 | 25 | 78 | 62 | 33 | 13 | 85 | | | | 18 |
| 8 | 10 | 93 | 25 | 78 | 62 | 33 | 13 | 19 | 77 | | | 19 |
| 8 | 10 | 93 | 25 | 78 | 62 | 33 | 13 | 19 | 38 | | | 20 |
| 7 | 10 | 93 | 25 | 78 | 62 | 33 | 13 | 41 | | | | 21 |
| 5 | 10 | 93 | 25 | 78 | 62 | 89 | | | | | | 22 |
| 7 | 10 | 93 | 25 | 78 | 62 | 18 | 35 | 36 | | | | 23 |
| 3 | 10 | 93 | 50 | 43 | | | | | | | | 24 |
| 153 | | | | | | | | | | | | |
| 6.375 | | | | | | | | | | | | |

Generations = 1481

| p=100 hr=4 mf=0.93 r0=1.88 inf=47 | | | | | | | | | | | | | | | | |
|---|---|---|---|---|---|---|---|---|---|---|---|---|---|---|---|---|
| Length | L0 | L1 | L2 | L3 | L4 | L5 | L6 | L7 | L8 | L9 | L10 | L11 | L12 | | | |
| 12 | 10 | 80 | 0 | 54 | 70 | 52 | 38 | 97 | 14 | 93 | 35 | 96 | 56 | | | 1 |
| 11 | 10 | 80 | 0 | 54 | 70 | 52 | 38 | 97 | 14 | 93 | 35 | 51 | | | | 2 |
| 10 | 10 | 80 | 0 | 54 | 70 | 52 | 38 | 97 | 14 | 93 | 20 | | | | | 3 |
| 10 | 10 | 80 | 0 | 54 | 70 | 52 | 38 | 97 | 14 | 93 | 64 | | | | | 4 |
| 11 | 10 | 80 | 0 | 54 | 70 | 52 | 38 | 97 | 14 | 46 | 31 | 91 | | | | 5 |
| 12 | 10 | 80 | 0 | 54 | 70 | 52 | 38 | 97 | 14 | 6 | 84 | 39 | 47 | | | 6 |
| 12 | 10 | 80 | 0 | 54 | 70 | 52 | 38 | 97 | 14 | 6 | 84 | 39 | 77 | | | 7 |
| 12 | 10 | 80 | 0 | 54 | 70 | 52 | 38 | 97 | 14 | 6 | 84 | 30 | 41 | | | 8 |
| 12 | 10 | 80 | 0 | 54 | 70 | 52 | 38 | 97 | 14 | 6 | 84 | 30 | 2 | | | 9 |
| 11 | 10 | 80 | 0 | 54 | 70 | 52 | 38 | 97 | 14 | 6 | 84 | 66 | | | | 10 |
| 10 | 10 | 80 | 0 | 54 | 70 | 52 | 38 | 97 | 14 | 6 | 87 | | | | | 11 |
| 10 | 10 | 80 | 0 | 54 | 70 | 52 | 38 | 97 | 14 | 6 | 34 | | | | | 12 |
| 7 | 10 | 80 | 0 | 54 | 70 | 52 | 38 | 85 | | | | | | | | 13 |
| 5 | 10 | 80 | 0 | 55 | 92 | 22 | | | | | | | | | | 14 |
| 7 | 10 | 80 | 0 | 55 | 92 | 83 | 68 | 60 | | | | | | | | 15 |
| 7 | 10 | 80 | 0 | 55 | 92 | 83 | 68 | 88 | | | | | | | | 16 |
| 7 | 10 | 80 | 0 | 55 | 92 | 83 | 68 | 5 | | | | | | | | 17 |
| 6 | 10 | 80 | 0 | 55 | 92 | 83 | 27 | | | | | | | | | 18 |
| 4 | 10 | 80 | 0 | 55 | 78 | | | | | | | | | | | 19 |
| 5 | 10 | 80 | 0 | 55 | 71 | 74 | | | | | | | | | | 20 |
| 4 | 10 | 80 | 16 | 28 | 9 | | | | | | | | | | | 21 |
| 2 | 10 | 13 | 19 | | | | | | | | | | | | | 22 |
| 187 | | | | | | | | | | | | | | | | |
| 8.5 | | | | | | | | | | | | | | | | |

Generations = 1748

| p=100 hr=4 mf=0.94 r0=2.12 inf=25 | | | | | | | | | | | | | | | | |
|---|---|---|---|---|---|---|---|---|---|---|---|---|---|---|---|---|
| Length | L0 | L1 | L2 | L3 | L4 | L5 | L6 | L7 | L8 | L9 | L10 | L11 | L12 | L13 | | |
| 4 | 10 | 23 | 13 | 42 | 54 | | | | | | | | | | | 1 |
| 3 | 10 | 23 | 62 | 60 | | | | | | | | | | | | 2 |
| 2 | 10 | 23 | 3 | | | | | | | | | | | | | 3 |
| 2 | 10 | 23 | 8 | | | | | | | | | | | | | 4 |
| 6 | 10 | 23 | 14 | 72 | 53 | 22 | 74 | | | | | | | | | 5 |
| 13 | 10 | 23 | 14 | 72 | 53 | 22 | 46 | 16 | 37 | 35 | 51 | 56 | 48 | 61 | | 6 |
| 13 | 10 | 23 | 14 | 72 | 53 | 22 | 46 | 16 | 37 | 35 | 51 | 56 | 48 | 95 | | 7 |
| 5 | 10 | 23 | 14 | 72 | 53 | 52 | | | | | | | | | | 8 |
| 5 | 10 | 23 | 14 | 72 | 53 | 49 | | | | | | | | | | 9 |
| 3 | 10 | 23 | 14 | 78 | | | | | | | | | | | | 10 |
| 56 | | | | | | | | | | | | | | | | |
| 5.6 | | | | | | | | | | | | | | | | |

Generations = 2588

| HR=4 NF=0.95 50new inf, R0=2.28 | | | | | | | | | | | | | | | | |
|---|---|---|---|---|---|---|---|---|---|---|---|---|---|---|---|---|
| Length | L0 | L1 | L2 | L3 | L4 | L5 | L6 | L7 | L8 | L9 | L10 | L11 | L12 | | | |
| 7 | 10 | 11 | 21 | 63 | 36 | 43 | 62 | 58 | | | | | | | | 1 |
| 4 | 10 | 11 | 21 | 63 | 72 | | | | | | | | | | | 2 |
| 4 | 10 | 11 | 21 | 23 | 32 | | | | | | | | | | | 3 |
| 3 | 10 | 11 | 73 | 8 | | | | | | | | | | | | 4 |
| 3 | 10 | 11 | 92 | 49 | | | | | | | | | | | | 5 |
| 3 | 10 | 11 | 92 | 42 | | | | | | | | | | | | 6 |
| 3 | 10 | 11 | 92 | 88 | | | | | | | | | | | | 7 |
| 3 | 10 | 11 | 92 | 69 | | | | | | | | | | | | 8 |
| 5 | 10 | 11 | 92 | 83 | 47 | 61 | | | | | | | | | | 9 |
| 5 | 10 | 11 | 92 | 83 | 26 | 5 | | | | | | | | | | 10 |
| 5 | 10 | 11 | 92 | 83 | 26 | 70 | | | | | | | | | | 11 |
| 5 | 10 | 11 | 92 | 83 | 16 | 71 | | | | | | | | | | 12 |
| 5 | 10 | 11 | 92 | 83 | 16 | 44 | | | | | | | | | | 13 |
| 4 | 10 | 11 | 3 | 14 | 34 | | | | | | | | | | | 14 |
| 3 | 10 | 11 | 3 | 37 | | | | | | | | | | | | 15 |
| 3 | 10 | 11 | 3 | 91 | | | | | | | | | | | | 16 |
| 5 | 10 | 11 | 84 | 33 | 66 | 75 | | | | | | | | | | 17 |
| 6 | 10 | 11 | 84 | 33 | 66 | 15 | 77 | | | | | | | | | 18 |
| 5 | 10 | 11 | 84 | 33 | 66 | 78 | | | | | | | | | | 19 |
| 2 | 10 | 11 | 76 | | | | | | | | | | | | | 20 |
| 2 | 10 | 18 | 99 | | | | | | | | | | | | | 21 |
| 2 | 10 | 13 | 22 | | | | | | | | | | | | | 22 |
| 6 | 10 | 13 | 45 | 31 | 0 | 81 | 20 | | | | | | | | | 23 |
| 3 | 10 | 13 | 48 | 4 | | | | | | | | | | | | 24 |
| 96 | | | | | | | | | | | | | | | | |
| 4.363636 | | | | | | | | | | | | | | | | |

Generations = 1056

| p=100 hr=4 mf=0.96 r=2.20 inf=50 Gen=1672 | | | | | | | | | | | | | | | | |
|---|---|---|---|---|---|---|---|---|---|---|---|---|---|---|---|---|
| Length | L0 | L1 | L2 | L3 | L4 | L5 | L6 | L7 | L8 | L9 | L10 | L11 | L12 | | | |
| 2 | 10 | 38 | 78 | | | | | | | | | | | | | 1 |
| 2 | 10 | 38 | 59 | | | | | | | | | | | | | 2 |
| 6 | 10 | 38 | 66 | 82 | 31 | 40 | 60 | | | | | | | | | 3 |
| 7 | 10 | 38 | 66 | 82 | 31 | 96 | 52 | 79 | | | | | | | | 4 |
| 5 | 10 | 38 | 66 | 82 | 57 | 81 | | | | | | | | | | 5 |
| 5 | 10 | 38 | 66 | 82 | 1 | 39 | | | | | | | | | | 6 |
| 6 | 10 | 38 | 66 | 85 | 76 | 8 | 18 | | | | | | | | | 7 |
| 7 | 10 | 38 | 66 | 85 | 53 | 13 | 20 | 55 | | | | | | | | 8 |
| 7 | 10 | 38 | 66 | 85 | 53 | 13 | 20 | 47 | | | | | | | | 9 |
| 11 | 10 | 38 | 66 | 85 | 53 | 13 | 20 | 64 | 24 | 45 | 15 | 4 | | | | 10 |
| 11 | 10 | 38 | 66 | 85 | 53 | 13 | 20 | 64 | 24 | 45 | 15 | 80 | | | | 11 |
| 10 | 10 | 38 | 66 | 85 | 53 | 13 | 20 | 64 | 24 | 45 | 86 | | | | | 12 |
| 10 | 10 | 38 | 66 | 85 | 53 | 13 | 20 | 64 | 98 | 72 | 34 | | | | | 13 |
| 10 | 10 | 38 | 66 | 85 | 53 | 13 | 20 | 64 | 98 | 72 | 37 | | | | | 14 |
| 9 | 10 | 38 | 66 | 85 | 53 | 13 | 20 | 64 | 98 | 0 | | | | | | 15 |
| 8 | 10 | 38 | 66 | 85 | 53 | 13 | 20 | 64 | 84 | | | | | | | 16 |
| 9 | 10 | 38 | 66 | 85 | 53 | 49 | 26 | 71 | 99 | 17 | | | | | | 17 |
| 9 | 10 | 38 | 66 | 85 | 53 | 49 | 26 | 71 | 99 | 6 | | | | | | 18 |
| 9 | 10 | 38 | 66 | 85 | 53 | 49 | 26 | 71 | 2 | 7 | | | | | | 19 |
| 8 | 10 | 38 | 66 | 85 | 53 | 49 | 26 | 71 | 54 | | | | | | | 20 |
| 6 | 10 | 38 | 66 | 85 | 53 | 49 | 91 | | | | | | | | | 21 |
| 6 | 10 | 38 | 66 | 85 | 53 | 49 | 9 | | | | | | | | | 22 |
| 5 | 10 | 38 | 66 | 85 | 75 | 41 | | | | | | | | | | 23 |
| 168 | | | | | | | | | | | | | | | | |
| 7.304348 | | | | | | | | | | | | | | | | |

| p=100 hr=4 mf=0.97 r0=2.70 inf=50 Gen=815 | | | | | | | | | | |
|---|---|---|---|---|---|---|---|---|---|---|
| Length | L0 | L1 | L2 | L3 | L4 | L5 | L6 | L7 | L8 | |
| 4 | 10 | 12 | 1 | 7 | 64 | | | | | 1 |
| 4 | 10 | 12 | 93 | 65 | 0 | | | | | 2 |
| 5 | 10 | 12 | 93 | 69 | 19 | 95 | | | | 3 |
| 5 | 10 | 12 | 93 | 69 | 19 | 92 | | | | 4 |
| 2 | 10 | 20 | 88 | | | | | | | 5 |
| 6 | 10 | 99 | 2 | 55 | 38 | 58 | 6 | | | 6 |
| 6 | 10 | 99 | 2 | 55 | 38 | 58 | 75 | | | 7 |
| 5 | 10 | 99 | 2 | 55 | 38 | 34 | | | | 8 |
| 5 | 10 | 99 | 2 | 41 | 87 | 43 | | | | 9 |
| 5 | 10 | 99 | 2 | 37 | 4 | 85 | | | | 10 |
| 5 | 10 | 99 | 2 | 37 | 4 | 68 | | | | 11 |
| 3 | 10 | 53 | 73 | 39 | | | | | | 12 |
| 5 | 10 | 53 | 62 | 22 | 15 | 11 | | | | 13 |
| 6 | 10 | 53 | 62 | 22 | 15 | 30 | 49 | | | 14 |
| 5 | 10 | 53 | 62 | 22 | 17 | 80 | | | | 15 |
| 5 | 10 | 53 | 62 | 22 | 17 | 24 | | | | 16 |
| 4 | 10 | 53 | 62 | 22 | 97 | | | | | 17 |
| 5 | 10 | 53 | 62 | 22 | 14 | 79 | | | | 18 |
| 3 | 10 | 53 | 62 | 50 | | | | | | 19 |
| 2 | 10 | 3 | 51 | | | | | | | 20 |
| 4 | 10 | 3 | 94 | 27 | 89 | | | | | 21 |
| 4 | 10 | 3 | 94 | 27 | 83 | | | | | 22 |
| 98 | | | | | | | | | | |
| 4.454545 | | | | | | | | | | |

| p=100 hr=4 mf=0.98 r0=2.40 inf=50 Gen=1602 | | | | | | | | | | | | | | | | |
|---|---|---|---|---|---|---|---|---|---|---|---|---|---|---|---|---|
| Length | L0 | L1 | L2 | L3 | L4 | L5 | L6 | L7 | L8 | L9 | L10 | L11 | L12 | | | |
| 9 | 10 | 62 | 11 | 79 | 24 | 82 | 25 | 48 | 58 | 19 | | | | | | 1 |
| 9 | 10 | 62 | 11 | 79 | 24 | 82 | 25 | 48 | 58 | 59 | | | | | | 2 |
| 11 | 10 | 62 | 11 | 79 | 24 | 82 | 25 | 48 | 89 | 47 | 6 | 93 | | | | 3 |
| 11 | 10 | 62 | 11 | 79 | 24 | 82 | 25 | 48 | 89 | 47 | 6 | 57 | | | | 4 |
| 11 | 10 | 62 | 11 | 79 | 24 | 82 | 25 | 48 | 89 | 47 | 6 | 66 | | | | 5 |
| 10 | 10 | 62 | 11 | 79 | 24 | 82 | 25 | 48 | 89 | 47 | 28 | | | | | 6 |
| 11 | 10 | 62 | 11 | 79 | 24 | 82 | 25 | 48 | 89 | 47 | 21 | 78 | | | | 7 |
| 11 | 10 | 62 | 11 | 79 | 24 | 82 | 25 | 48 | 89 | 47 | 21 | 35 | | | | 8 |
| 11 | 10 | 62 | 11 | 79 | 24 | 82 | 25 | 48 | 89 | 47 | 21 | 73 | | | | 9 |
| 10 | 10 | 62 | 11 | 79 | 24 | 82 | 25 | 48 | 89 | 47 | 87 | | | | | 10 |
| 9 | 10 | 62 | 11 | 79 | 24 | 82 | 25 | 48 | 89 | 71 | | | | | | 11 |
| 7 | 10 | 62 | 11 | 79 | 24 | 82 | 25 | 43 | | | | | | | | 12 |
| 8 | 10 | 62 | 11 | 79 | 24 | 82 | 25 | 46 | 45 | | | | | | | 13 |
| 7 | 10 | 62 | 11 | 79 | 24 | 82 | 25 | 94 | | | | | | | | 14 |
| 3 | 10 | 62 | 11 | 91 | | | | | | | | | | | | 15 |
| 3 | 10 | 62 | 7 | 9 | | | | | | | | | | | | 16 |
| 3 | 10 | 62 | 7 | 60 | | | | | | | | | | | | 17 |
| 3 | 10 | 62 | 7 | 39 | | | | | | | | | | | | 18 |
| 3 | 10 | 62 | 83 | 0 | | | | | | | | | | | | 19 |
| 7 | 10 | 62 | 80 | 99 | 44 | 30 | 97 | 75 | | | | | | | | 20 |
| 6 | 10 | 62 | 80 | 99 | 44 | 30 | 86 | | | | | | | | | 21 |
| 3 | 10 | 62 | 38 | 72 | | | | | | | | | | | | 22 |
| 3 | 10 | 62 | 27 | 20 | | | | | | | | | | | | 23 |
| 3 | 10 | 62 | 27 | 51 | | | | | | | | | | | | 24 |
| 4 | 10 | 62 | 84 | 1 | 15 | | | | | | | | | | | 25 |
| 4 | 10 | 62 | 84 | 1 | 55 | | | | | | | | | | | 26 |
| 180 | | | | | | | | | | | | | | | | |
| 6.923077 | | | | | | | | | | | | | | | | |

| p=100 hr=4 mf=0.99 r0=2.06 inf=49 d76h18 Gen=1531 | | | | | | | | | | | | | | | | |
|---|---|---|---|---|---|---|---|---|---|---|---|---|---|---|---|---|
| Length | L0 | L1 | L2 | L3 | L4 | L5 | L6 | L7 | L8 | L9 | L10 | L11 | L12 | | | |
| 3 | 10 | 54 | 64 | 18 | | | | | | | | | | | | 1 |
| 7 | 10 | 54 | 64 | 44 | 41 | 82 | 71 | 69 | | | | | | | | 2 |
| 8 | 10 | 54 | 64 | 44 | 41 | 82 | 71 | 27 | 13 | | | | | | | 3 |
| 9 | 10 | 54 | 64 | 44 | 41 | 82 | 71 | 27 | 73 | 32 | | | | | | 4 |
| 6 | 10 | 54 | 64 | 44 | 41 | 48 | 2 | | | | | | | | | 5 |
| 8 | 10 | 54 | 64 | 44 | 41 | 48 | 92 | 21 | 16 | | | | | | | 6 |
| 5 | 10 | 54 | 64 | 44 | 98 | 20 | | | | | | | | | | 7 |
| 4 | 10 | 54 | 64 | 99 | 49 | | | | | | | | | | | 8 |
| 6 | 10 | 28 | 72 | 34 | 79 | 76 | 0 | | | | | | | | | 9 |
| 5 | 10 | 28 | 72 | 34 | 51 | 94 | | | | | | | | | | 10 |
| 5 | 10 | 28 | 72 | 34 | 51 | 9 | | | | | | | | | | 11 |
| 7 | 10 | 28 | 72 | 30 | 70 | 91 | 53 | 39 | | | | | | | | 12 |
| 9 | 10 | 28 | 72 | 30 | 70 | 91 | 11 | 40 | 81 | 12 | | | | | | 13 |
| 9 | 10 | 28 | 72 | 30 | 70 | 91 | 11 | 40 | 81 | 62 | | | | | | 14 |
| 7 | 10 | 28 | 72 | 30 | 70 | 91 | 11 | 31 | | | | | | | | 15 |
| 7 | 10 | 28 | 72 | 30 | 70 | 91 | 11 | 97 | | | | | | | | 16 |
| 8 | 10 | 28 | 72 | 30 | 70 | 91 | 11 | 23 | 65 | | | | | | | 17 |
| 8 | 10 | 28 | 72 | 30 | 70 | 91 | 11 | 68 | 61 | | | | | | | 18 |
| 6 | 10 | 28 | 72 | 30 | 70 | 91 | 15 | | | | | | | | | 19 |
| 4 | 10 | 28 | 72 | 36 | 59 | | | | | | | | | | | 20 |
| 131 | | | | | | | | | | | | | | | | |
| 6.55 | | | | | | | | | | | | | | | | |

| P=100 Hr=4 mf=1.00 R0=3.68 inf=50 Gen=924 | | | | | | | | | | | | | | |
|---|---|---|---|---|---|---|---|---|---|---|---|---|---|---|
| Length | L0 | L1 | L2 | L3 | L4 | L5 | L6 | L7 | L8 | L9 | L10 | L11 | L12 | |
| 2 | 10 | 87 | 60 | | | | | | | | | | | 1 |
| 5 | 10 | 87 | 22 | 61 | 20 | 34 | | | | | | | | 2 |
| 5 | 10 | 87 | 22 | 68 | 53 | 16 | | | | | | | | 3 |
| 5 | 10 | 87 | 22 | 68 | 51 | 89 | | | | | | | | 4 |
| 5 | 10 | 87 | 22 | 68 | 51 | 25 | | | | | | | | 5 |
| 5 | 10 | 87 | 92 | 8 | 43 | 46 | | | | | | | | 6 |
| 6 | 10 | 87 | 92 | 8 | 43 | 55 | 97 | | | | | | | 7 |
| 5 | 10 | 87 | 92 | 67 | 83 | | | | | | | | | 8 |
| 5 | 10 | 87 | 92 | 67 | 33 | | | | | | | | | 9 |
| 7 | 10 | 87 | 92 | 67 | 26 | 54 | 27 | 3 | | | | | | 10 |
| 7 | 10 | 87 | 92 | 67 | 26 | 54 | 27 | 47 | | | | | | 11 |
| 6 | 10 | 87 | 92 | 67 | 26 | 54 | 32 | | | | | | | 12 |
| 6 | 10 | 87 | 92 | 67 | 26 | 54 | 71 | | | | | | | 13 |
| 6 | 10 | 87 | 92 | 67 | 26 | 82 | 93 | | | | | | | 14 |
| 6 | 10 | 87 | 92 | 67 | 26 | 82 | 65 | | | | | | | 15 |
| 5 | 10 | 87 | 92 | 67 | 59 | 84 | | | | | | | | 16 |
| 5 | 10 | 87 | 92 | 67 | 59 | 63 | | | | | | | | 17 |
| 5 | 10 | 87 | 92 | 67 | 59 | 73 | | | | | | | | 18 |
| 5 | 10 | 87 | 92 | 67 | 59 | 30 | | | | | | | | 19 |
| 5 | 10 | 87 | 92 | 67 | 59 | 75 | | | | | | | | 20 |
| 2 | 10 | 63 | 85 | | | | | | | | | | | 21 |
| 5 | 10 | 6 | 90 | 39 | 86 | 38 | | | | | | | | 22 |
| 5 | 10 | 6 | 90 | 39 | 86 | 1 | | | | | | | | 23 |
| 7 | 10 | 6 | 90 | 66 | 18 | 0 | 79 | 99 | | | | | | 24 |
| 6 | 10 | 6 | 90 | 66 | 18 | 0 | 40 | | | | | | | 25 |
| 2 | 10 | 6 | 72 | | | | | | | | | | | 26 |
| 133 | | | | | | | | | | | | | | |
| 5.115 | Average Depth | | | | | | | | | | | | | |

| p=100 hr=4 mf=1.01 r0=1.96 inf=47 d56h3 Gen=1056 | | | | | | | | | | | | | | | | |
|---|---|---|---|---|---|---|---|---|---|---|---|---|---|---|---|---|
| Length | L0 | L1 | L2 | L3 | L4 | L5 | L6 | L7 | L8 | L9 | L10 | L11 | L12 | | | |
| 6 | 10 | 37 | 82 | 47 | 39 | 95 | 87 | | | | | | | | | 1 |
| 5 | 10 | 37 | 82 | 44 | 60 | 54 | | | | | | | | | | 2 |
| 6 | 10 | 37 | 82 | 44 | 9 | 93 | 85 | | | | | | | | | 3 |
| 7 | 10 | 37 | 82 | 44 | 9 | 51 | 11 | 49 | | | | | | | | 4 |
| 5 | 10 | 37 | 82 | 44 | 9 | 0 | | | | | | | | | | 5 |
| 5 | 10 | 37 | 82 | 44 | 9 | 34 | | | | | | | | | | 6 |
| 6 | 10 | 37 | 82 | 44 | 16 | 38 | 30 | | | | | | | | | 7 |
| 4 | 10 | 37 | 82 | 44 | 29 | | | | | | | | | | | 8 |
| 5 | 10 | 37 | 82 | 44 | 22 | 23 | | | | | | | | | | 9 |
| 5 | 10 | 78 | 3 | 58 | 41 | 12 | | | | | | | | | | 10 |
| 4 | 10 | 78 | 3 | 6 | 33 | | | | | | | | | | | 11 |
| 5 | 10 | 78 | 3 | 6 | 15 | 45 | | | | | | | | | | 12 |
| 6 | 10 | 78 | 3 | 6 | 15 | 84 | 98 | | | | | | | | | 13 |
| 5 | 10 | 78 | 3 | 6 | 57 | 1 | | | | | | | | | | 14 |
| 6 | 10 | 78 | 3 | 6 | 57 | 70 | 96 | | | | | | | | | 15 |
| 6 | 10 | 78 | 3 | 6 | 7 | 83 | 40 | | | | | | | | | 16 |
| 6 | 10 | 78 | 3 | 6 | 7 | 83 | 19 | | | | | | | | | 17 |
| 5 | 10 | 78 | 3 | 6 | 7 | 64 | | | | | | | | | | 18 |
| 5 | 10 | 78 | 3 | 6 | 7 | 53 | | | | | | | | | | 19 |
| 5 | 10 | 78 | 3 | 6 | 7 | 81 | | | | | | | | | | 20 |
| 5 | 10 | 78 | 3 | 6 | 72 | 65 | | | | | | | | | | 21 |
| 112 | | | | | | | | | | | | | | | | |
| 5.3333 | | | | | | | | | | | | | | | | |

| p=100 hr=4 mf=1.02 r0=2.79 inf=50 50:4:4:13:29 Gen=1238 | | | | | | | | | | | | |
|---|---|---|---|---|---|---|---|---|---|---|---|---|
| Length | L0 | L1 | L2 | L3 | L4 | L5 | L6 | L7 | L8 | l9 | | |
| 3 | 10 | 87 | 65 | 23 | | | | | | | | 1 |
| 7 | 10 | 87 | 65 | 18 | 79 | 55 | 67 | 3 | | | | 2 |
| 8 | 10 | 87 | 65 | 18 | 79 | 55 | 67 | 74 | 78 | | | 3 |
| 8 | 10 | 87 | 65 | 18 | 79 | 55 | 67 | 74 | 31 | | | 4 |
| 5 | 10 | 87 | 65 | 18 | 79 | 34 | | | | | | 5 |
| 6 | 10 | 87 | 65 | 18 | 79 | 98 | 15 | | | | | 6 |
| 4 | 10 | 87 | 65 | 18 | 44 | | | | | | | 7 |
| 7 | 10 | 87 | 96 | 64 | 81 | 26 | 36 | 21 | | | | 8 |
| 8 | 10 | 87 | 96 | 64 | 81 | 62 | 22 | 20 | 38 | | | 9 |
| 9 | 10 | 87 | 96 | 64 | 81 | 62 | 22 | 20 | 12 | 93 | | 10 |
| 9 | 10 | 87 | 96 | 64 | 81 | 62 | 22 | 0 | 68 | 14 | | 11 |
| 9 | 10 | 87 | 96 | 64 | 81 | 62 | 22 | 0 | 68 | 33 | | 12 |
| 8 | 10 | 87 | 96 | 64 | 81 | 62 | 22 | 0 | 1 | | | 13 |
| 8 | 10 | 87 | 96 | 64 | 81 | 62 | 22 | 11 | 27 | | | 14 |
| 8 | 10 | 87 | 96 | 64 | 81 | 62 | 22 | 11 | 76 | | | 15 |
| 8 | 10 | 87 | 96 | 64 | 81 | 62 | 22 | 11 | 39 | | | 16 |
| 7 | 10 | 87 | 96 | 64 | 81 | 62 | 22 | 61 | | | | 17 |
| 8 | 10 | 87 | 96 | 64 | 81 | 62 | 2 | 50 | 63 | | | 18 |
| 8 | 10 | 87 | 96 | 64 | 81 | 62 | 2 | 50 | 48 | | | 19 |
| 8 | 10 | 87 | 96 | 64 | 81 | 62 | 2 | 50 | 53 | | | 20 |
| 7 | 10 | 87 | 96 | 64 | 81 | 62 | 2 | 29 | | | | 21 |
| 5 | 10 | 87 | 96 | 64 | 81 | 72 | | | | | | 22 |
| 5 | 10 | 87 | 96 | 64 | 81 | 43 | | | | | | 23 |
| 5 | 10 | 87 | 49 | 25 | 45 | 54 | | | | | | 24 |
| 4 | 10 | 87 | 49 | 25 | 8 | | | | | | | 25 |
| 172 | | | | | | | | | | | | |
| 6.88 | | | | | | | | | | | | |

| p=100 hr=4 mf=1.03 r0=2.25 inf=50 49:4:0:7:40 Gen=1017 | | | | | | | | | | |
|---|---|---|---|---|---|---|---|---|---|---|
| Length | L0 | L1 | L2 | L3 | L4 | L5 | L6 | L7 | L8 | |
| 5 | 10 | 96 | 94 | 55 | 31 | 66 | | | | 1 |
| 6 | 10 | 96 | 94 | 55 | 31 | 30 | 26 | | | 2 |
| 8 | 10 | 96 | 94 | 55 | 31 | 30 | 27 | 64 | 43 | 3 |
| 6 | 10 | 96 | 94 | 55 | 31 | 30 | 71 | | | 4 |
| 5 | 10 | 96 | 94 | 55 | 31 | 4 | | | | 5 |
| 3 | 10 | 96 | 94 | 35 | | | | | | 6 |
| 6 | 10 | 96 | 90 | 99 | 74 | 41 | 47 | | | 7 |
| 6 | 10 | 96 | 90 | 99 | 74 | 50 | 45 | | | 8 |
| 5 | 10 | 96 | 90 | 99 | 74 | 75 | | | | 9 |
| 6 | 10 | 96 | 90 | 99 | 3 | 79 | 87 | | | 10 |
| 6 | 10 | 96 | 90 | 99 | 3 | 82 | 14 | | | 11 |
| 7 | 10 | 96 | 90 | 99 | 3 | 82 | 98 | 24 | | 12 |
| 7 | 10 | 96 | 90 | 99 | 3 | 82 | 98 | 13 | | 13 |
| 7 | 10 | 96 | 90 | 99 | 3 | 82 | 98 | 34 | | 14 |
| 4 | 10 | 96 | 90 | 99 | 15 | | | | | 15 |
| 4 | 10 | 96 | 90 | 99 | 25 | | | | | 16 |
| 4 | 10 | 96 | 90 | 29 | 61 | | | | | 17 |
| 4 | 10 | 96 | 90 | 29 | 81 | | | | | 18 |
| 4 | 10 | 28 | 48 | 22 | 72 | | | | | 19 |
| 4 | 10 | 28 | 48 | 93 | 59 | | | | | 20 |
| 3 | 10 | 28 | 67 | 17 | | | | | | 21 |
| 3 | 10 | 28 | 51 | 52 | | | | | | 22 |
| 3 | 10 | 28 | 9 | 88 | | | | | | 23 |
| 2 | 10 | 91 | 6 | | | | | | | 24 |
| 2 | 10 | 91 | 62 | | | | | | | 25 |
| 120 | | | | | | | | | | |
| 4.8 | | | | | | | | | | |

| p=100 hr=4 mf=1.04 r0=3.41 50:4:9:15:22Gen=855 | | | | | | | | | |
|---|---|---|---|---|---|---|---|---|---|
| Length | L0 | L1 | L2 | L3 | L4 | L5 | L6 | L7 | L8 |
| 1 | 10 | 85 | | | | | | | 1 |
| 2 | 10 | 43 | 49 | | | | | | 2 |
| 3 | 10 | 43 | 81 | 25 | | | | | 3 |
| 4 | 10 | 90 | 76 | 46 | 54 | | | | 4 |
| 3 | 10 | 90 | 76 | 20 | | | | | 5 |
| 4 | 10 | 90 | 76 | 11 | 14 | | | | 6 |
| 4 | 10 | 90 | 76 | 11 | 88 | | | | 7 |
| 5 | 10 | 90 | 76 | 11 | 36 | 6 | | | 8 |
| 4 | 10 | 90 | 76 | 11 | 94 | | | | 9 |
| 4 | 10 | 90 | 76 | 11 | 2 | | | | 10 |
| 5 | 10 | 90 | 76 | 11 | 28 | 5 | | | 11 |
| 4 | 10 | 90 | 40 | 72 | 77 | | | | 12 |
| 3 | 10 | 90 | 40 | 16 | | | | | 13 |
| 4 | 10 | 31 | 12 | 21 | 71 | | | | 14 |
| 3 | 10 | 31 | 12 | 64 | | | | | 15 |
| 2 | 10 | 31 | 50 | | | | | | 16 |
| 6 | 10 | 31 | 89 | 13 | 7 | 60 | 32 | | 17 |
| 4 | 10 | 31 | 89 | 13 | 8 | | | | 18 |
| 6 | 10 | 31 | 89 | 44 | 35 | 42 | 22 | | 19 |
| 6 | 10 | 31 | 89 | 44 | 35 | 42 | 15 | | 20 |
| 6 | 10 | 31 | 89 | 44 | 35 | 42 | 75 | | 21 |
| 6 | 10 | 31 | 89 | 44 | 35 | 42 | 38 | | 22 |
| 6 | 10 | 31 | 89 | 44 | 35 | 4 | 73 | | 23 |
| 5 | 10 | 31 | 89 | 44 | 35 | 62 | | | 24 |
| 5 | 10 | 31 | 89 | 44 | 35 | 87 | | | 25 |
| 5 | 10 | 31 | 89 | 44 | 35 | 74 | | | 26 |
| 4 | 10 | 31 | 89 | 44 | 30 | | | | 27 |
| 3 | 10 | 31 | 89 | 29 | | | | | 28 |
| 2 | 10 | 31 | 66 | | | | | | 29 |
| 119 | | | | | | | | | |
| 4.103 | | | | | | | | | |

| p=100 HzR=5 mF=2 Gen=568 | | | | | | | | |
|---|---|---|---|---|---|---|---|---|
| Length | L0 | L1 | L2 | L3 | L4 | L5 | L6 | |
| 5 | 10 | 52 | 58 | 51 | 40 | | | 1 |
| 4 | 10 | 52 | 66 | 45 | | | | 2 |
| 3 | 10 | 30 | 1 | | | | | 3 |
| 5 | 10 | 30 | 92 | 3 | 67 | | | 4 |
| 3 | 10 | 30 | 75 | | | | | 5 |
| 6 | 10 | 30 | 55 | 41 | 59 | 0 | | 6 |
| 5 | 10 | 30 | 55 | 56 | 48 | | | 7 |
| 5 | 10 | 30 | 74 | 7 | 97 | | | 8 |
| 4 | 10 | 30 | 82 | 12 | | | | 9 |
| 5 | 10 | 30 | 82 | 84 | 43 | | | 10 |
| 4 | 10 | 30 | 82 | 29 | | | | 11 |
| 4 | 10 | 30 | 37 | 26 | | | | 12 |
| 4 | 10 | 30 | 37 | 53 | | | | 13 |
| 2 | 10 | 71 | | | | | | 14 |
| 5 | 10 | 6 | 60 | 88 | 80 | | | 15 |
| 5 | 10 | 6 | 60 | 88 | 39 | | | 16 |
| 5 | 10 | 6 | 60 | 86 | 95 | | | 17 |
| 4 | 10 | 6 | 60 | 69 | | | | 18 |
| 4 | 10 | 6 | 21 | 17 | | | | 19 |
| 5 | 10 | 57 | 54 | 42 | 65 | | | 20 |
| 5 | 10 | 57 | 54 | 93 | 61 | | | 21 |
| 4 | 10 | 57 | 54 | 13 | | | | 22 |
| 3 | 10 | 36 | 47 | | | | | 23 |
| 99 | | | | | | | | |
| 4.304348 | | | | | | | | |
